# Use of Diagnosis Codes to Find Blood Transfusion Adverse Events in Electronic Health Records

**DOI:** 10.1101/2020.12.30.20218610

**Authors:** Roselie A. Bright, Susan J. Bright-Ponte, Lee Anne Palmer, Summer K. Rankin, Sergey Blok

## Abstract

**Background:** Electronic health records (EHRs) and big data tools offer the opportunity for surveillance of adverse events (patient harm associated with medical care). We chose the case of transfusion adverse events (TAEs) and potential TAEs (PTAEs) because 1.) real dates were obscured in the study data, and 2.) there was emerging recognition of new types during the study data period.

**Objective:** We aimed to use the structured data in electronic health records (EHRs) to find TAEs and PTAEs among adults.

**Methods:** We used 49,331 adult admissions involving critical care at a major teaching hospital, 2001-2012, in the MIMIC-III EHRs database. We formed a T (defined as packed red blood cells, platelets, or plasma) group of 21,443 admissions vs. 25,468 comparison (C) admissions. The ICD-9-CM diagnosis codes were compared for T vs. C, described, and tested with statistical tools.

**Results:** TAEs such as transfusion associated circulatory overload (TACO; 12 T cases; rate ratio (RR) 15.61; 95% CI 2.49 to 98) were found. There were also PTAEs similar to TAEs, such as fluid overload disorder (361 T admissions; RR 2.24; 95% CI 1.88 to 2.65), similar to TACO. Some diagnoses could have been sequelae of TAEs, including nontraumatic compartment syndrome of abdomen (52 T cases; RR 6.76; 95% CI 3.40 to 14.9) possibly being a consequence of TACO.

**Conclusions:** Surveillance for diagnosis codes that could be TAE sequelae or unrecognized TAE might be useful supplements to existing medical product adverse event programs.

## INTRODUCTION

FDA relies on spontaneous reports of patient harm (known as adverse events (AE)) from manufacturers, healthcare providers, and the general public to learn of AEs that may need regulatory action [1]. Published deficiencies of these reports [2-10] include varying representativeness of harm and problems. Now that electronic healthcare records (EHRs) are very common [11], we have an opportunity to leverage them as sources of AE information [4, 12-14].

Challenges to the automated use of diagnosis codes for surveillance include:

- ICD [International Classification of Diseases] codes may be “invalid, insensitive or non-specific” [15].
- EHRs are “…subject to access restrictions…” [2].
- “…[N]ot all events and outcomes are consistently captured …” [13].

The most directly useful program would analyze EHRs [1, 6] in real time. The data available to us (critical care admissions) did not allow calendar time analyses, so we compared blood transfusion admissions (T) vs. comparison admissions (C) because:

- An earlier version of the dataset showed a higher risk of near-term mortality for red blood cell transfusion patients, compared to non-transfused patients [16].
- During the time of the dataset, new transfusion AE (TAE) types, transfusion related acute lung injury (TRALI) and transfusion associated cardiac overload (TACO), were being recognized among transfusion experts and prompted new guidelines to reduce the use of blood transfusion [17].
- During the dataset time, far fewer reports were coming to FDA than would have been expected, considering the level of professional concern [18-20].

The focus of this study is on diagnosis codes in electronic health records (EHR) because it is a relatively available source of information in EHRs and are readily computable. Many studies have looked for predetermined unwelcome conditions; for example, emergency department visits due to opioid overdose [21], motor vehicle crash injury [22], and serious infections [23]. We studied the utility of this method in this setting so that we can compare the results to using another surveillance method with the same data.

The objective of this study was to observe the utility of diagnosis codes to find TAEs that could be further evaluated (outside this study’s scope) with a formal analysis to demonstrate causality.

## METHODS

We used EHRs for critical care admissions within an adult hospital, Beth Israel Deaconess Medical Center, Boston, MA. The Massachusetts Institute of Technology (MIT) worked with the hospital to process EHRs, including unstructured notes, from 2001 to 2012 into a privacy-protected dataset, Medical Information Mart for Intensive Care III (MIMIC-III) [24, 25], that is publicly available to those meeting human subjects research requirements. The research was designated not human subjects research by the FDA Institutional Review Board under Code of Federal Regulations Title 45 Part 46 [26].

We removed admissions with patient age < 16 years on the date of admission, as well as admissions without any notes, from the total of 58,976 admissions, resulting in 49,284 admissions.

The transfused group (T) of 21,443 admissions had at least one transfusion (red blood cells, platelets, or plasma) “input code” in the “Inputs” table or International Statistical Classification of Diseases and Related Health Problems (ICD) 9-CM Volume 3 procedure code in the “Procedures” table, listed in Appendix Table A1 (Codes for transfusion and exclusion groups). Among the remaining patients, we identified 2,373 admissions whose only indication of possible transfusion are the codes listed in Appendix Table A1 and excluded them from further study. The other 25,468 admissions formed the comparison (C) group.

We counted the numbers of admissions in T and C for all ICD9 diagnoses [27] that occurred at least as often in T as in C, using Python packages SciPy Stats [28] and Pandas [29]. Most of the codes were present in the MIMIC-III dictionary of ICD9 diagnosis codes. For those that were not, names were found on a public website and enclosed in brackets [30]. When the choice of appropriate code name was not clear, we examined the procedure codes for the relevant admissions, and if necessary, the discharge notes to adjudicate the correct name.

We manually reviewed the information that was available for admissions that received an explicit TAE code despite being assigned to C.

For each diagnosis code we calculated the rate difference (RD), rate ratio (RR), their normal approximate 95% lower and upper confidence limits (95% LCL and 95% UCL), and p values [31]. When any of the rate numerators had a value of 5 or less, we calculated the Fishers Exact Test (FET) p value with Python and then derived the confidence limits [32].

Highly related codes were presented adjacent to each other in Tables 2 through 9 and subtotaled. All 524 diagnoses in the Tables met one of the following criteria [33-86]:

- Known TAE
- Hypercoagulation, embolism, or anemia diagnosis
- RR > 2
- RR > 1.5 and actual (T minus C) counts over 100
- The FET p value is < 0.05 (calculated when either of the T or C counts < 6)

TAE codes in our study were compared to TAE codes in published literature with a Fisher’s Exact Test calculator [87].

Our presentation of exact p values as low as “<0.0001” allows the reader to calculate their own adjustment of alpha based on their clinical interpretation of the interdependence of the diagnosis codes. If all 524 presented diagnoses are completely dependent, the alpha level to use is 0.0000979 [88]. Specific adjusted alpha levels assuming complete dependence are given with the results in tables organized by TAE-oriented system: iatrogenic (Table 2), circulation (3), bleeding and anemia (4), immune, neoplasm, and infection (5), lungs and oral (6), fluids regulation (7), digestion and hormones (8), and connective, nervous and reproductive (9).

We used published studies of TAEs in Medicare data [19, 20, 89, 90] to calculate expected TAE rates in MIMIC-III.

## RESULTS

Table 1 shows the explanations for all admissions that were assigned to the C group and had an explicit TAE diagnosis code. For six of the twelve admissions, the notes say transfusion occurred. For four, the notes say there was transfusion just before admission to the hospital or CCU. For the remaining two, there was no mention of transfusion in the notes, so the TAE diagnosis codes were not explained.

**Table 1.**
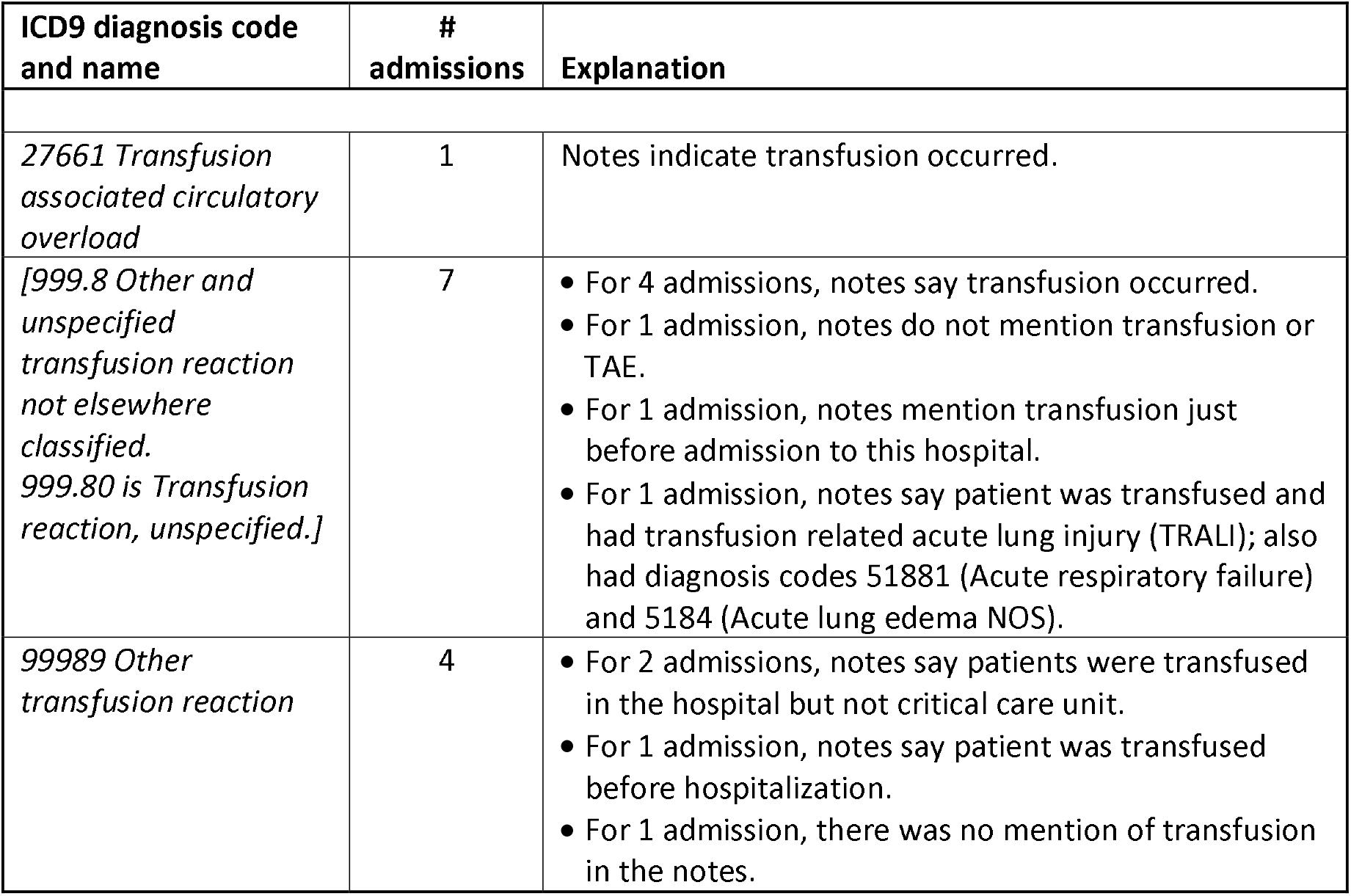
Explanations for all admissions that were assigned to the comparison group and had an explicit transfusion adverse event (TAE) diagnosis code (International Statistical Classification of Diseases and Related Health Problems 9-CM). Bracketed diagnosis code names were not in the MIMIC-III database, therefore were looked up [30] and confirmed in the notes.

| ICD9 diagnosis code and name | # admissions | Explanation |
| --- | --- | --- |
| 27661 *Transfusion associated circulatory overload* | 1 | Notes indicate transfusion occurred. |
| [999.8 *Other and unspecified transfusion reaction not elsewhere classified. 999.80 is Transfusion reaction, unspecified.*] | 7 | • For 4 admissions, notes say transfusion occurred.<br>• For 1 admission, notes do not mention transfusion or TAE.<br>• For 1 admission, notes mention transfusion just before admission to this hospital.<br>• For 1 admission, notes say patient was transfused and had transfusion related acute lung injury (TRALI); also had diagnosis codes 51881 (Acute respiratory failure) and 5184 (Acute lung edema NOS). |
| 99989 *Other transfusion reaction* | 4 | • For 2 admissions, notes say patients were transfused in the hospital but not critical care unit.<br>• For 1 admission, notes say patient was transfused before hospitalization.<br>• For 1 admission, there was no mention of transfusion in the notes. |

**Table 2.**
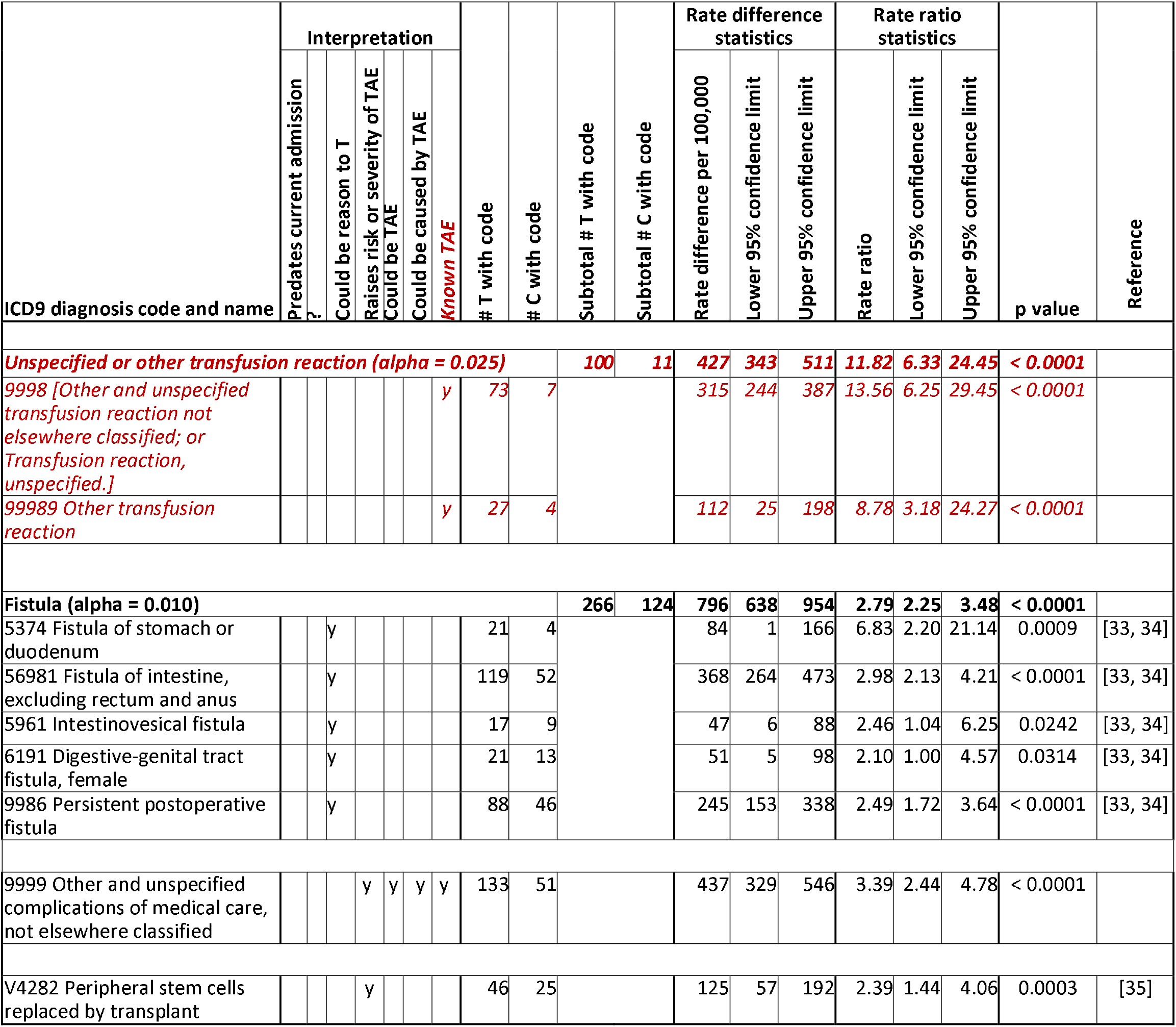
Analysis of T (transfusion) and C (comparison) groups for iatrogenic diagnosis codes. If all diagnoses in the table are dependent, alpha is 0.0057. Bracketed diagnosis code names were not in the MIMIC-III database, therefore were looked up [30] and confirmed in some notes.

The remaining tables have 524 diagnosis codes. The following TAE codes were found in the T group:

- ICD9 code 27661, Transfusion associated circulatory overload, in Table 7
- ICD9 code 27951, Acute graft-versus-host disease, in Table 5, which could apply to transplants or transfusion, in Table 5
- ICD9 code 78066, Febrile nonhemolytic transfusion reaction, in Table 5
- ICD9 code 9998, Other and unspecified transfusion reaction not elsewhere classified, in Table 2
- ICD9 code 99989, Other transfusion reaction, in Table 2

Most of them were at least one of: predated admission (53), known reasons or associated with reasons to transfuse (315), known risk factors for TAE (12), sequelae of TAE (120), clinically very similar to known TAE (120), or known TAE (13).

For two explicit TAE codes, codes for similar conditions were also statistically significantly more common in T.

- For febrile nonhemolytic transfusion reaction in Table 5, 78061 (Fever presenting with conditions classified elsewhere) was also much more common in T than C and was assigned much more often (196 admissions vs 3).
- For transfusion associated circulatory overload in Table 7, 2766 (fluid overload disorder) was also much more common in T than C and was assigned much more often (571 admissions vs 13).

**Table 3.**
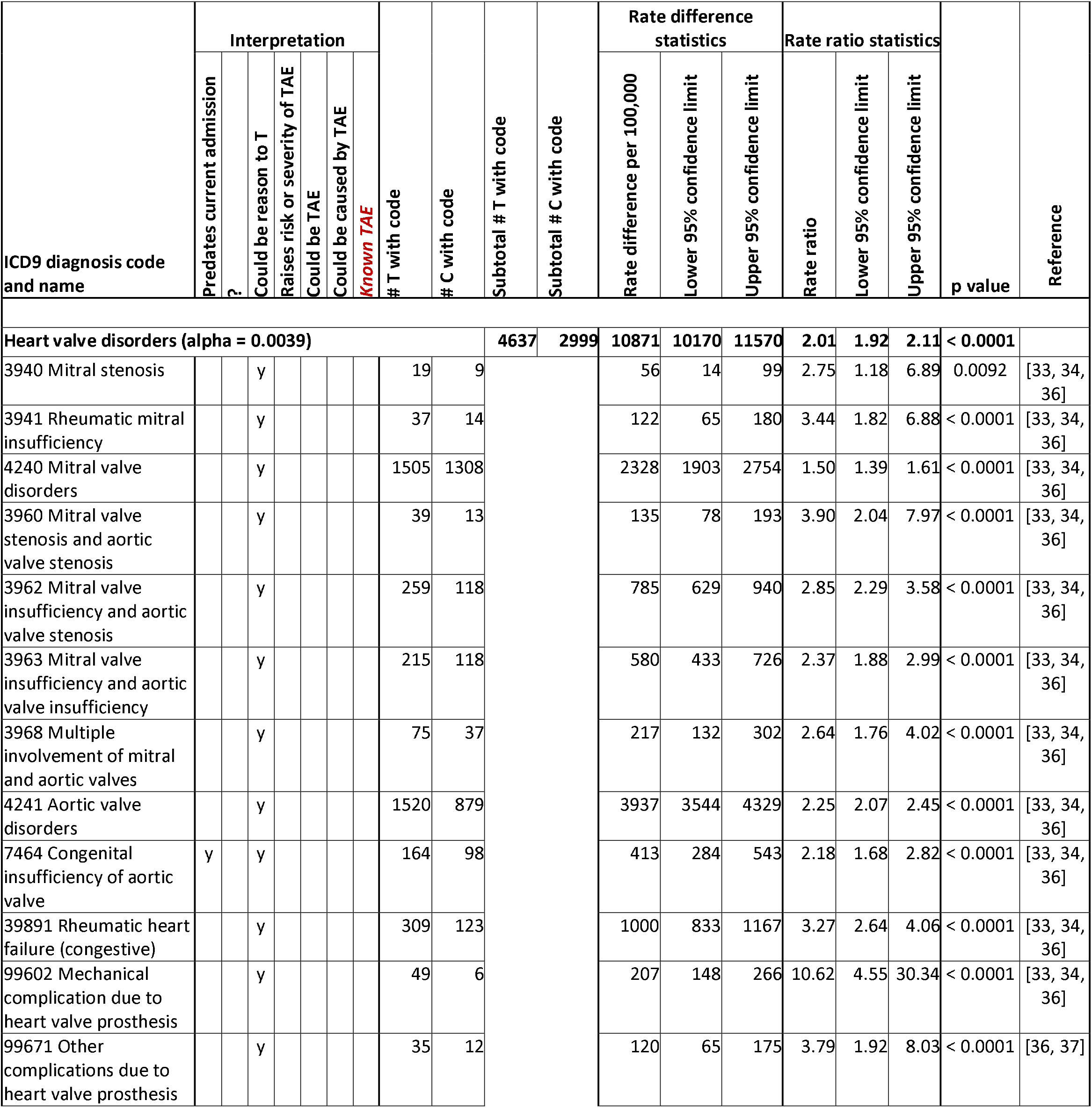
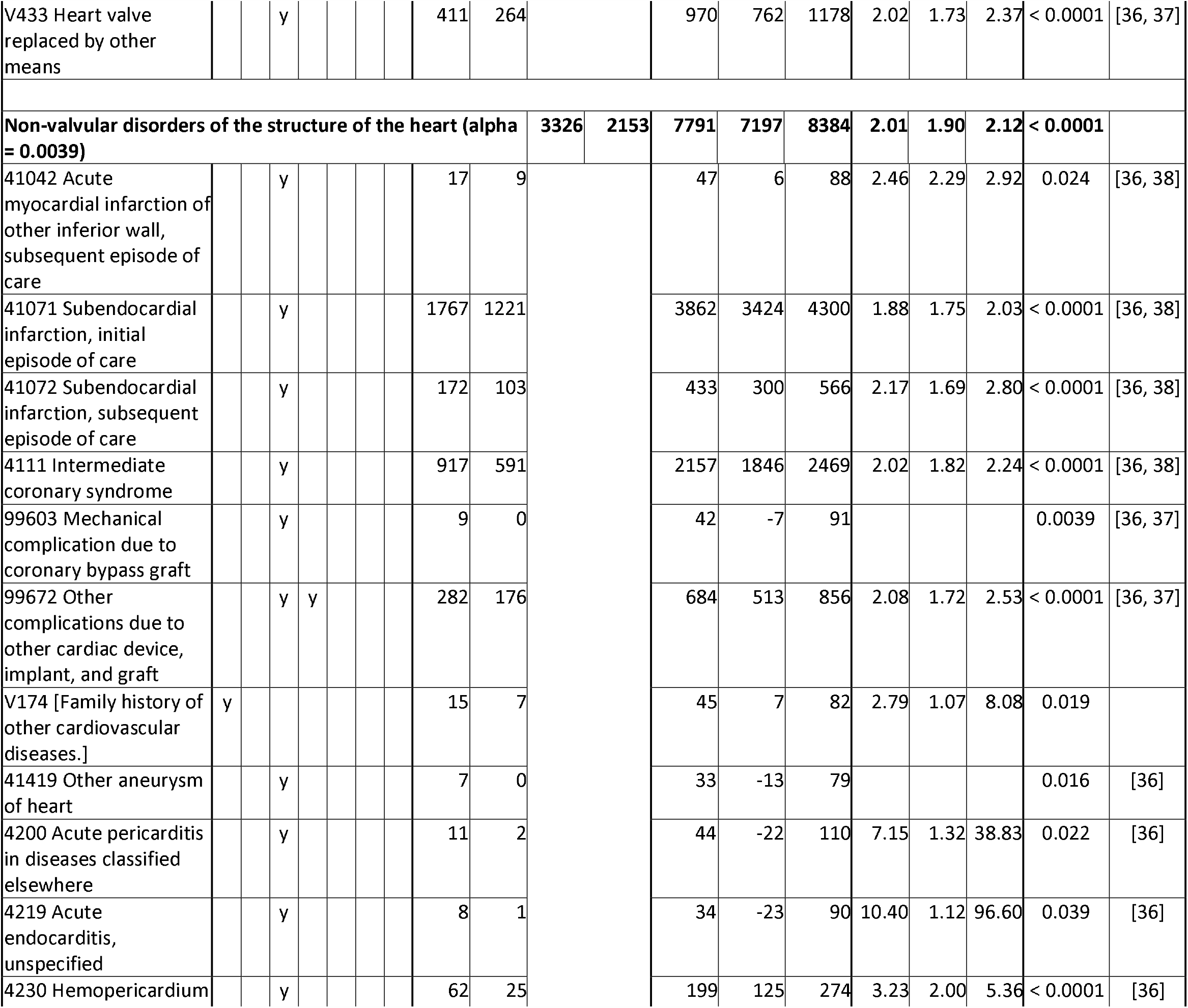
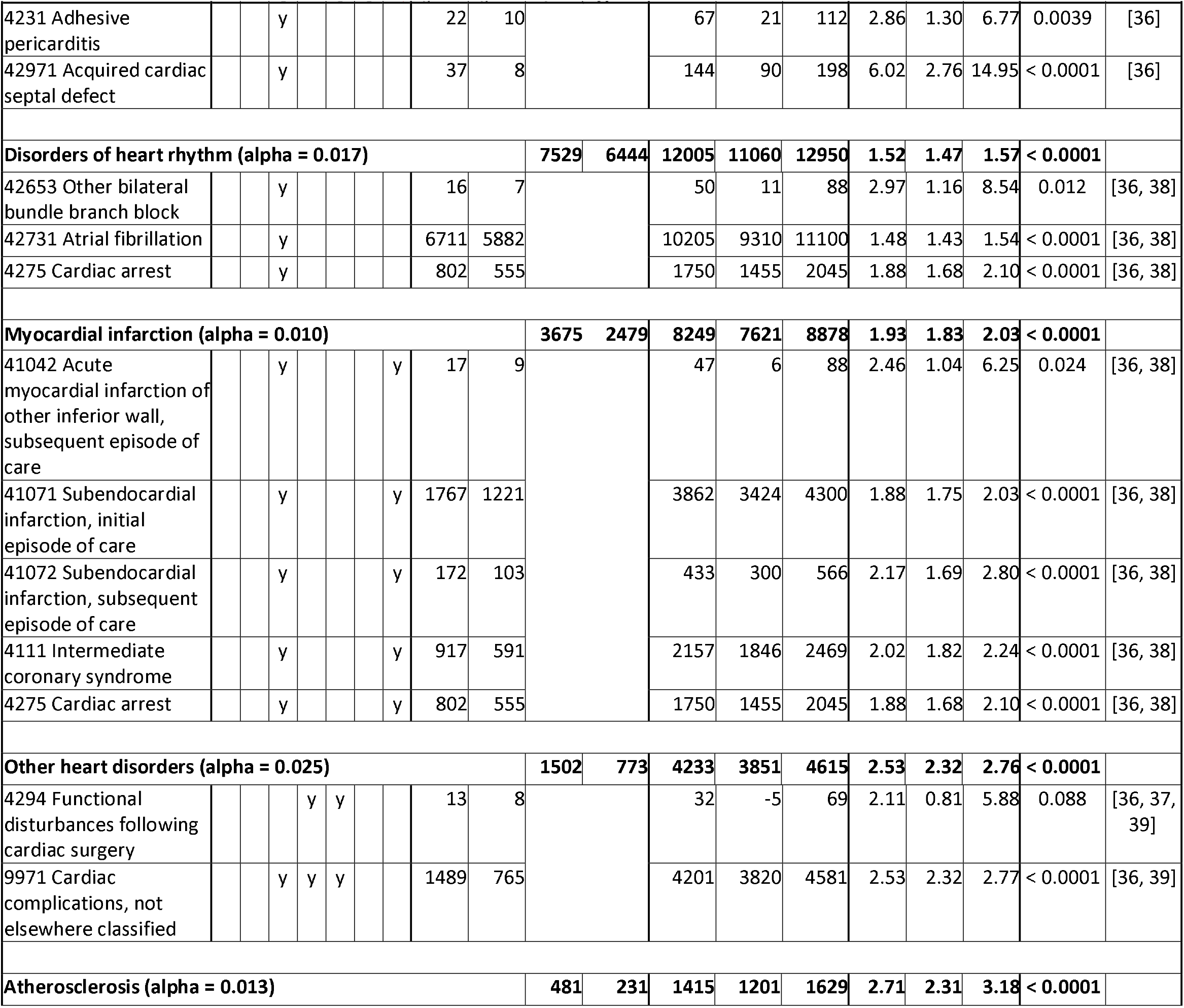
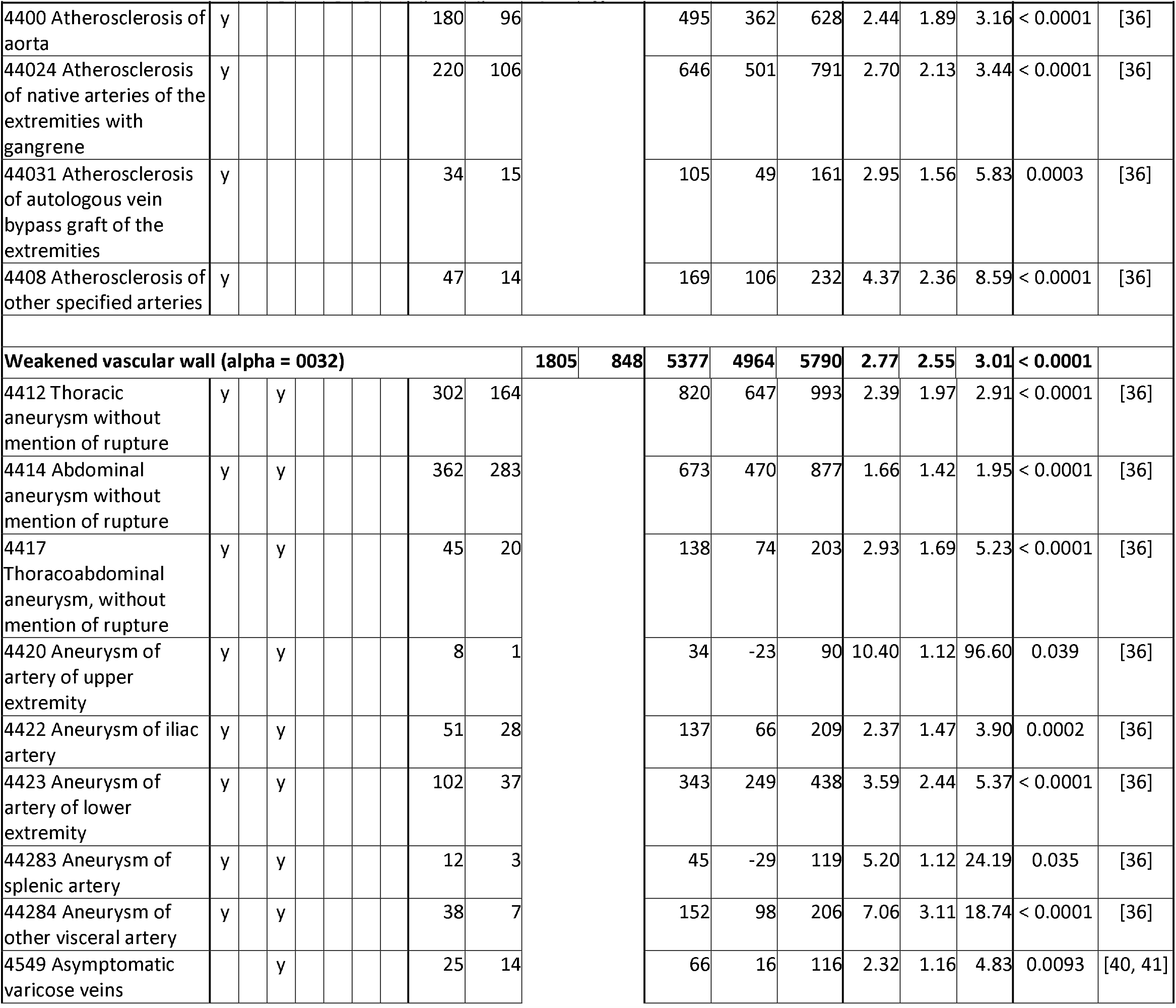
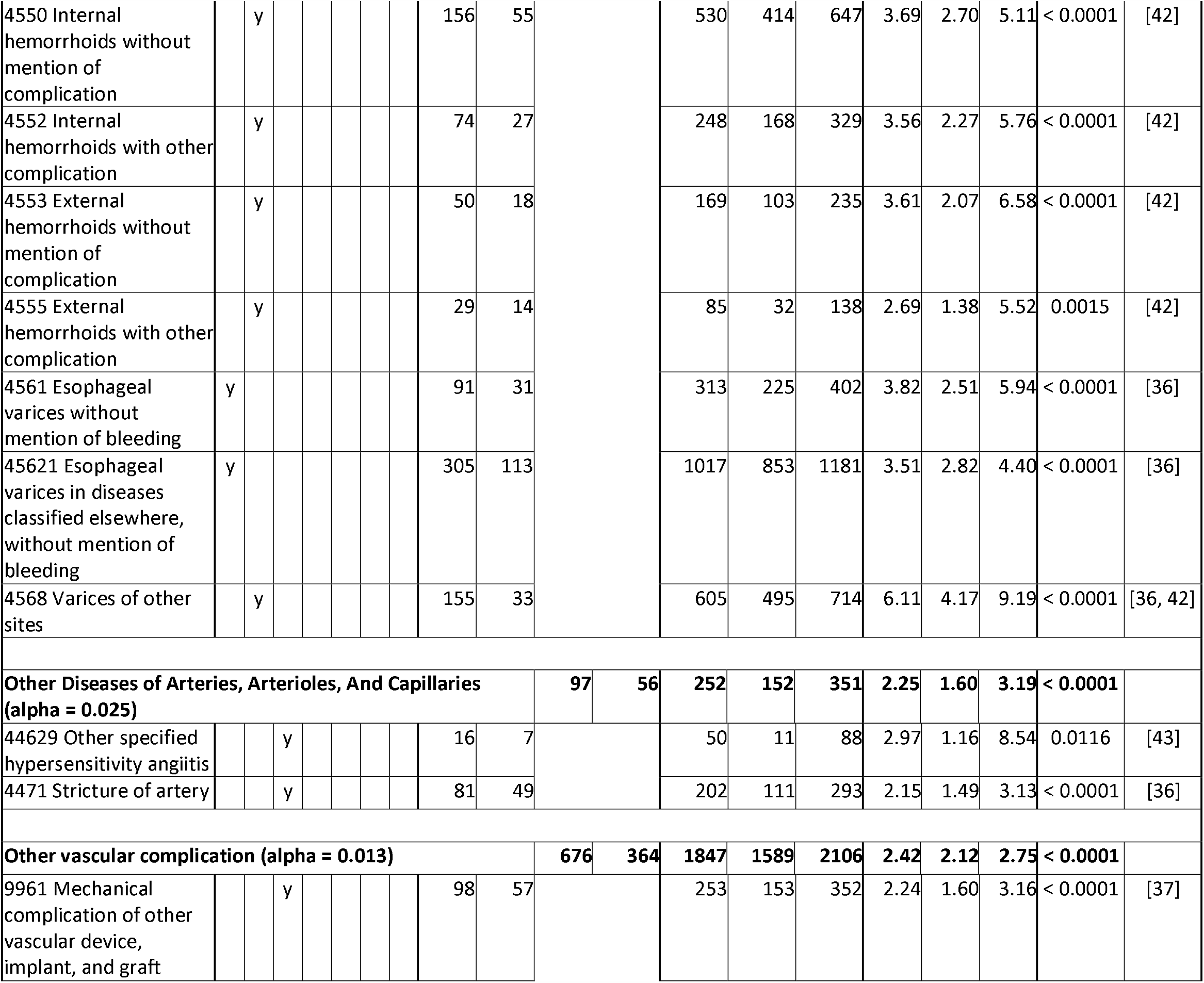
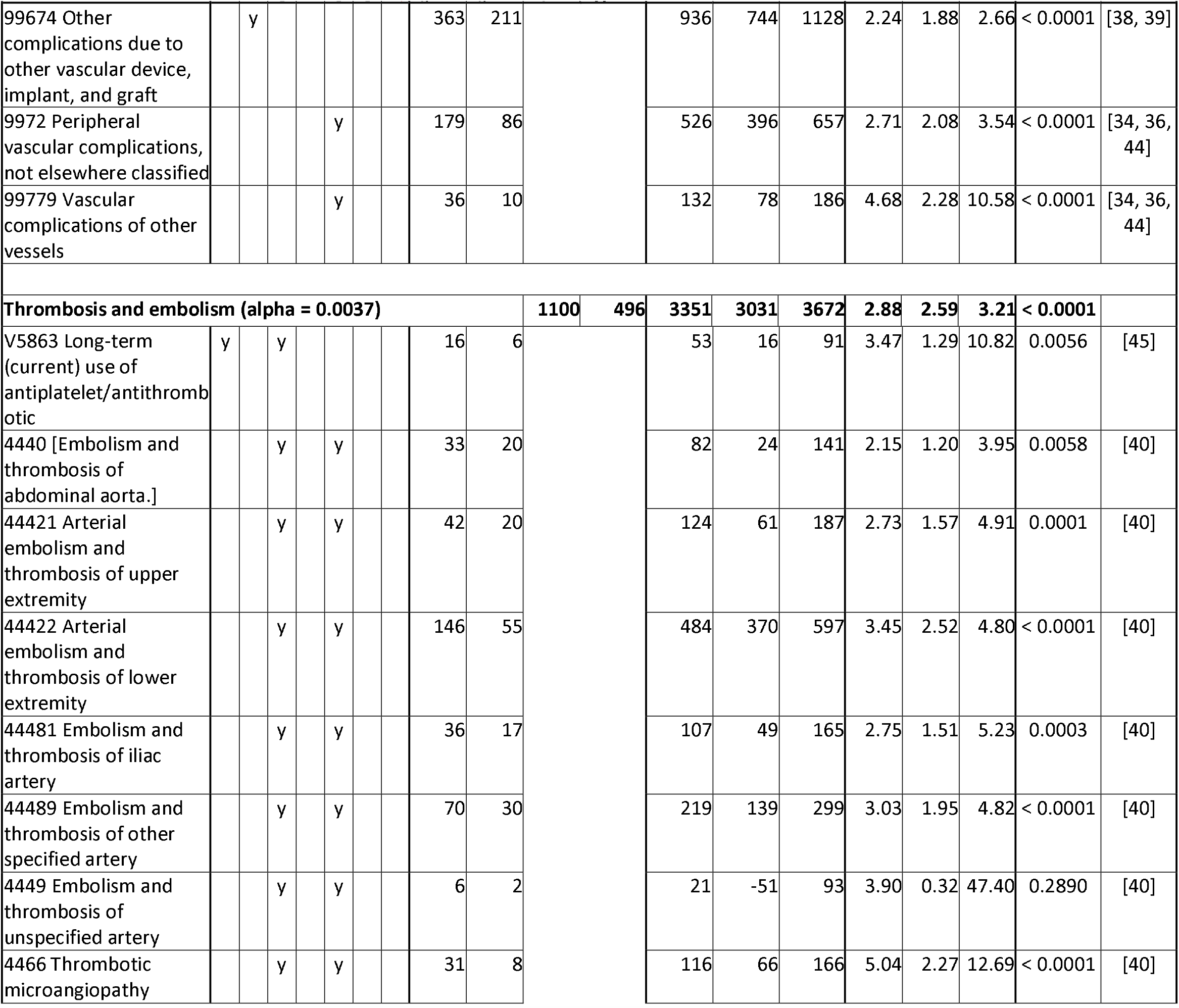
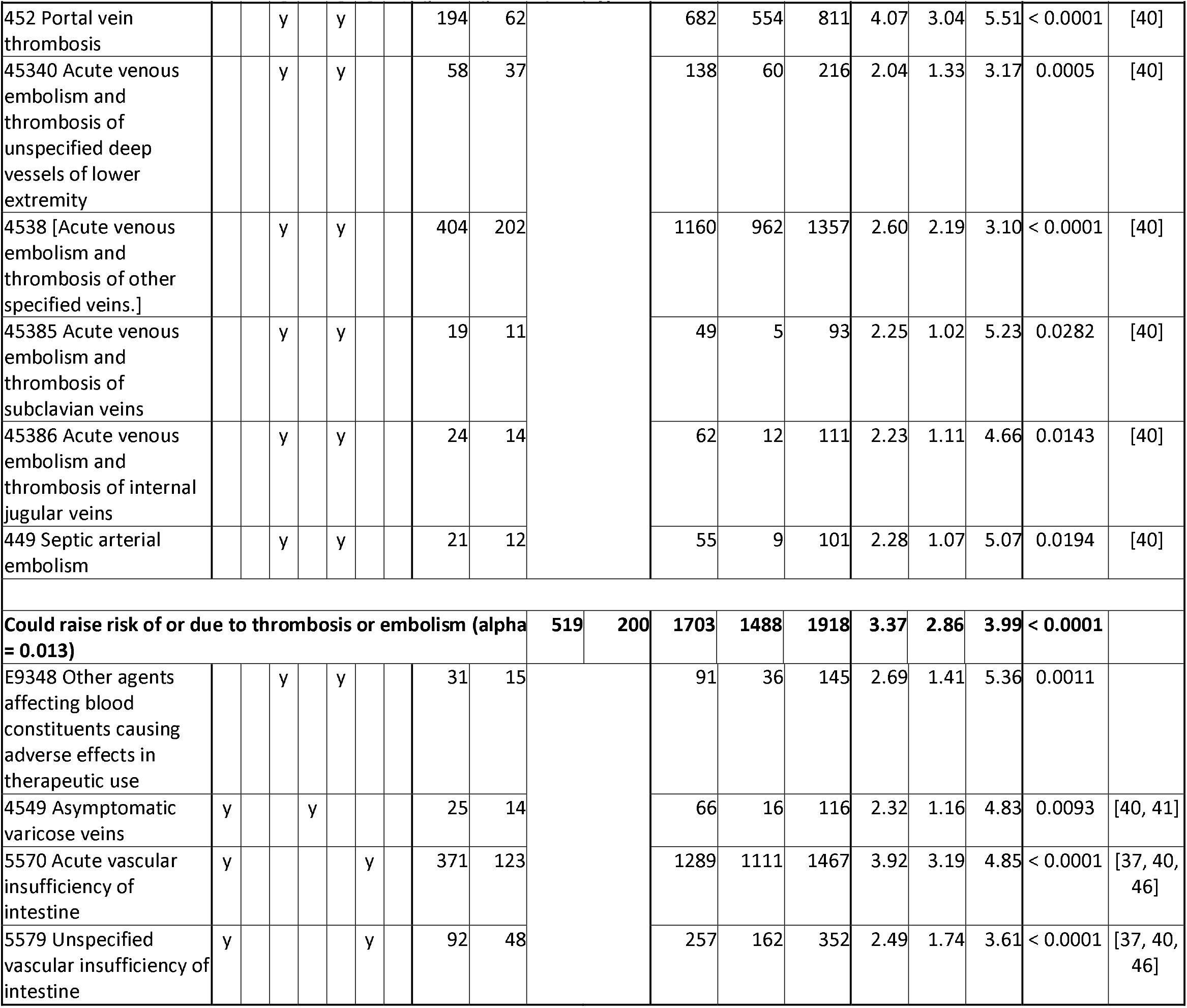
Analysis of T (transfusion) and C (comparison) groups for circulation diagnosis codes. If all diagnoses in the table are dependent, alpha is 0.00069. Bracketed diagnosis code names were not in the MIMIC-III database, therefore were looked up [30] and confirmed in some notes.

**Table 4.**
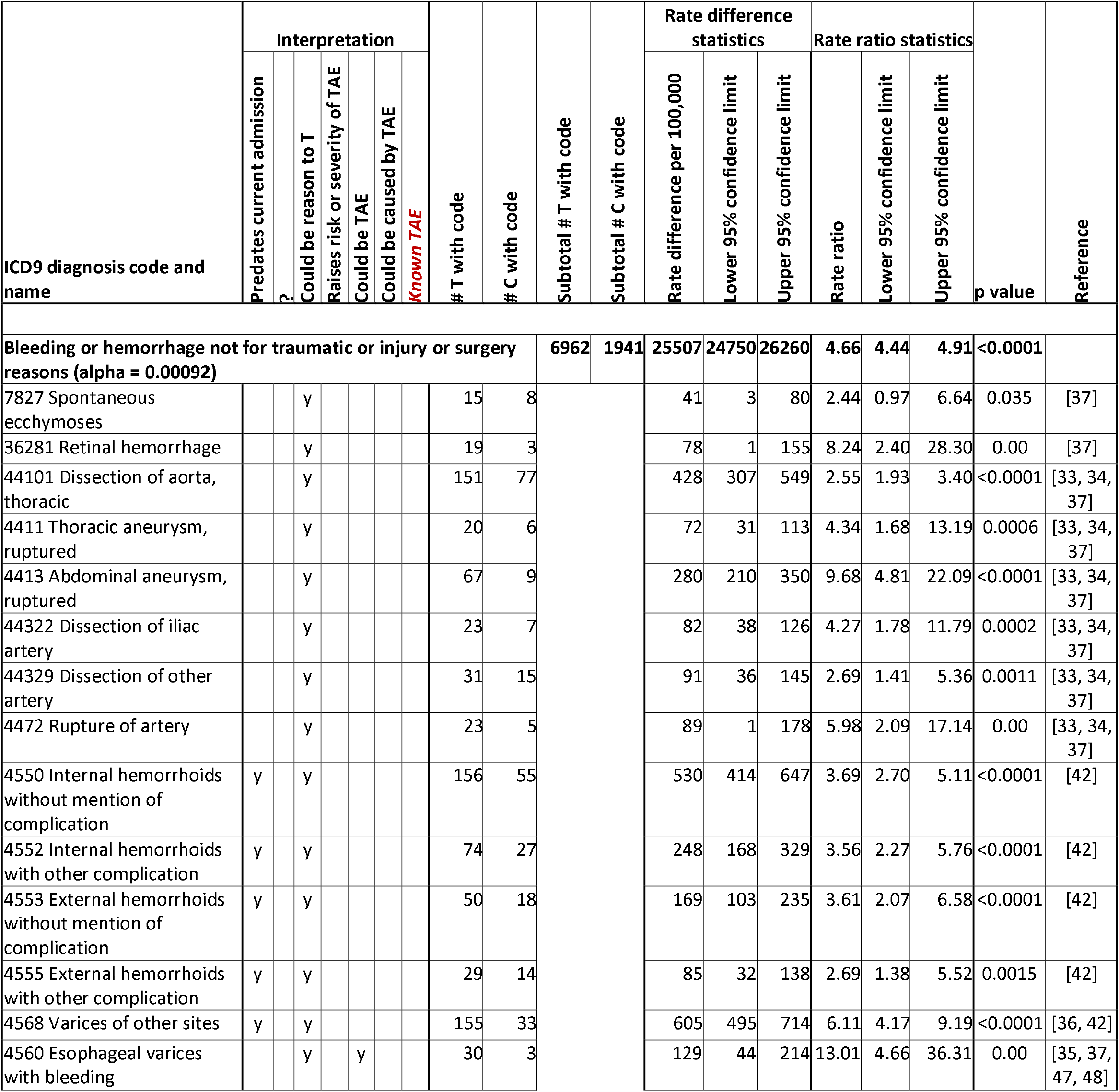
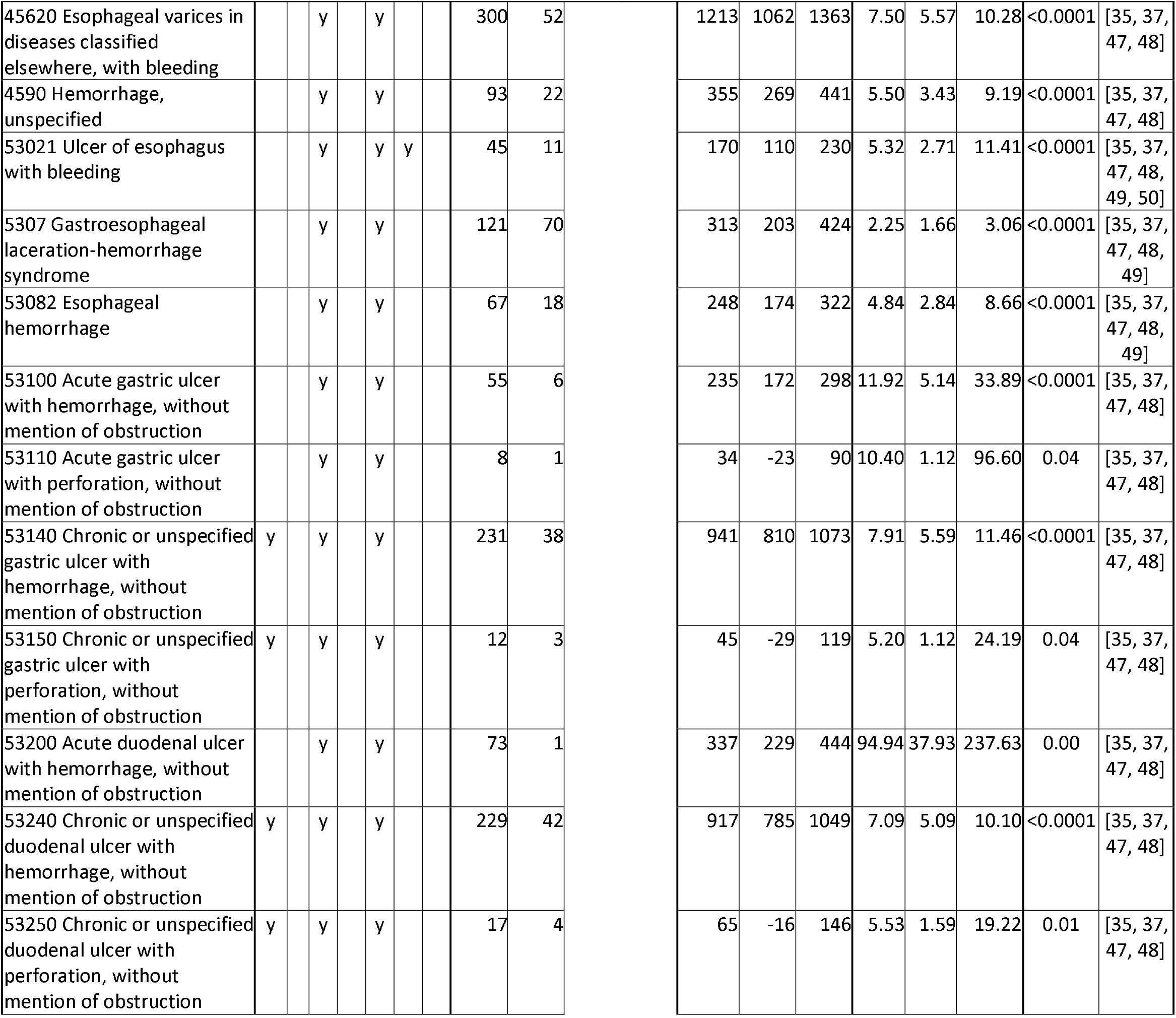
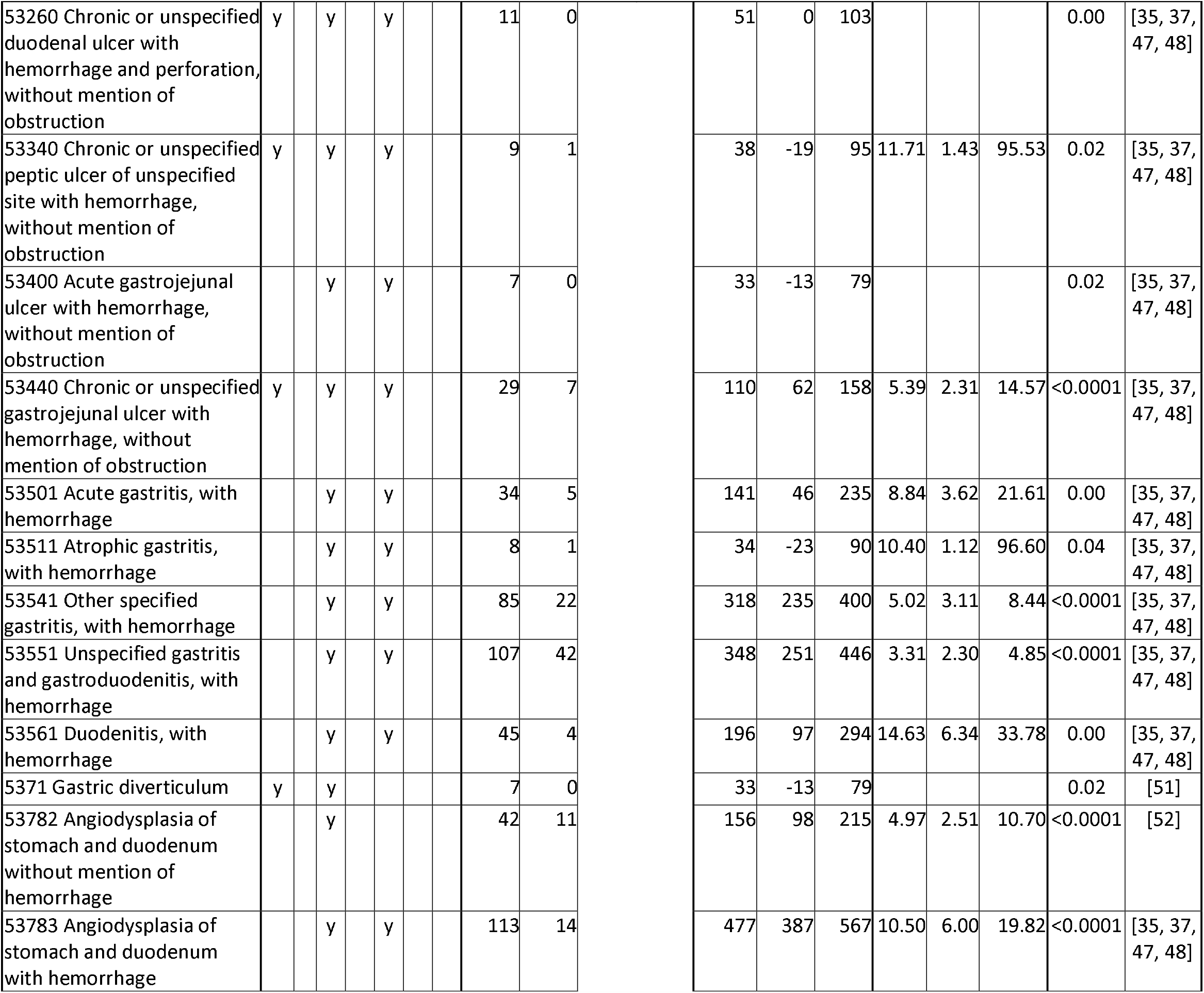
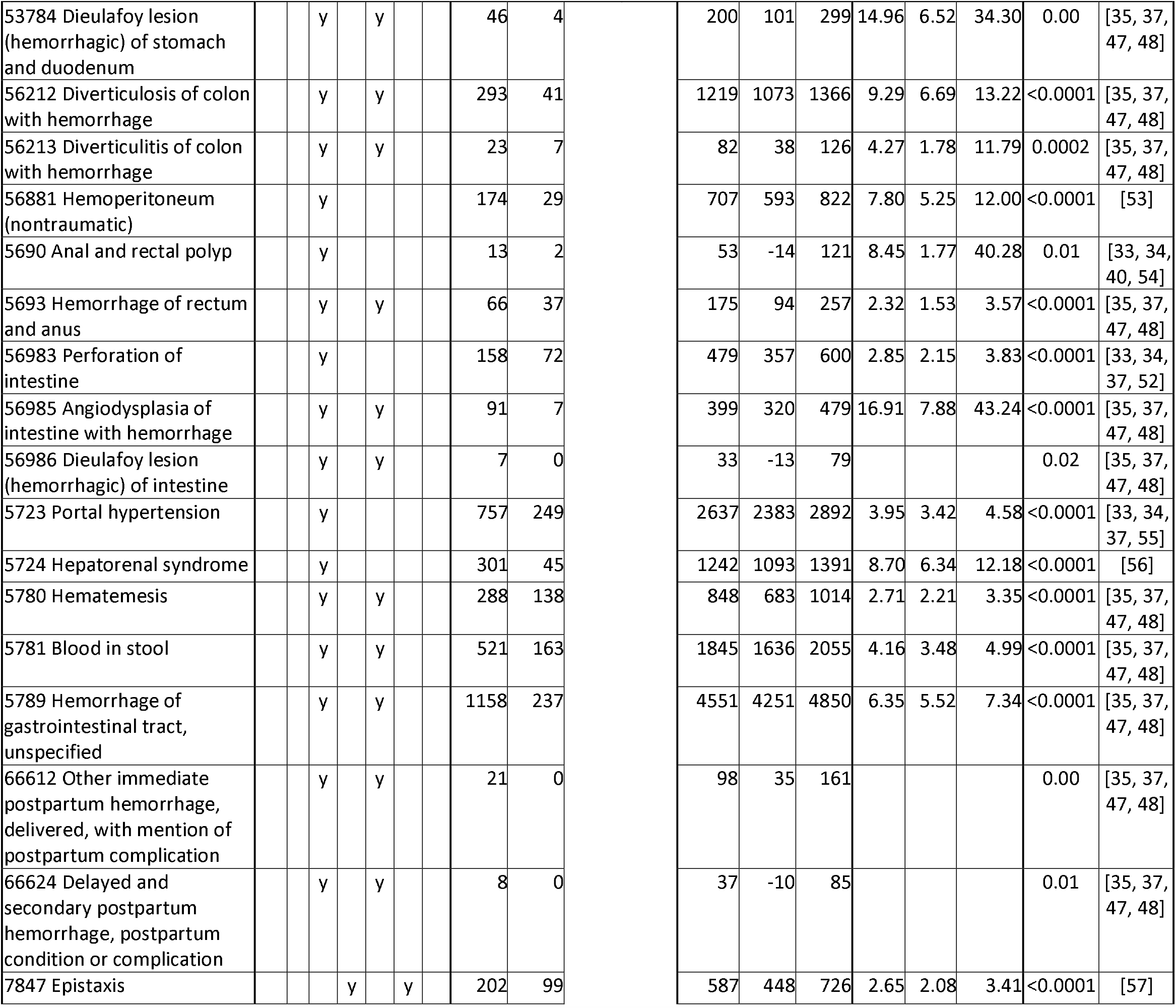
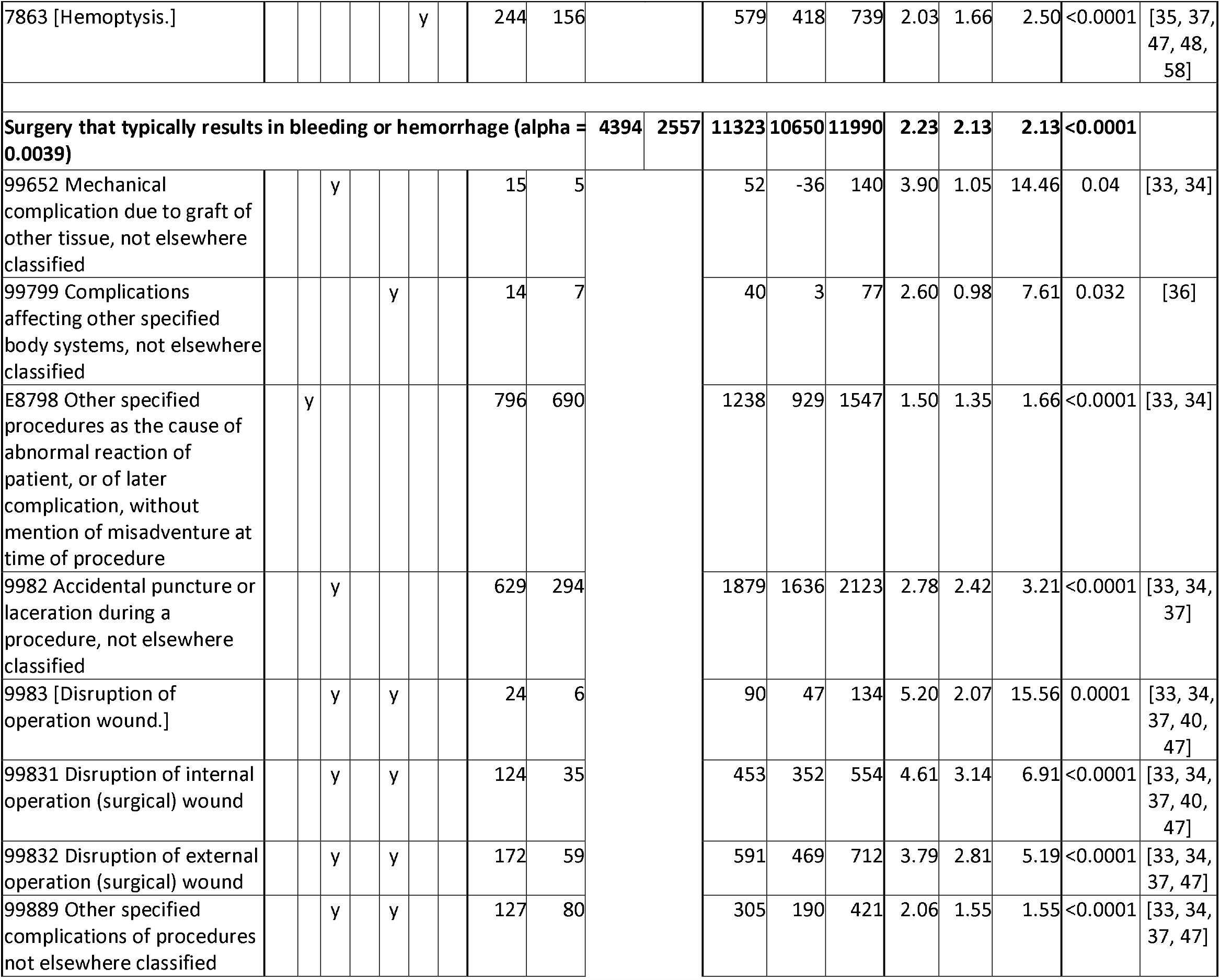
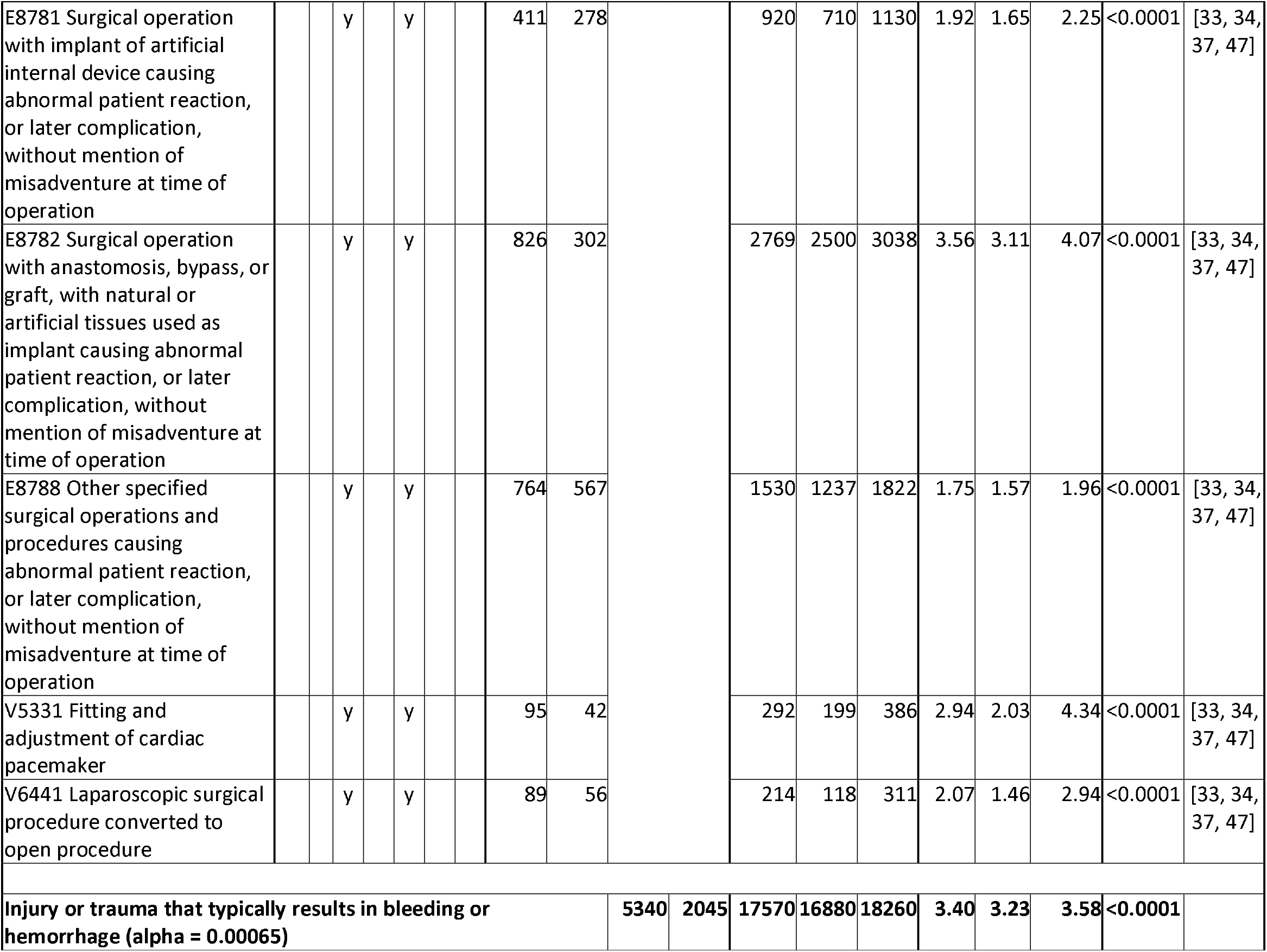
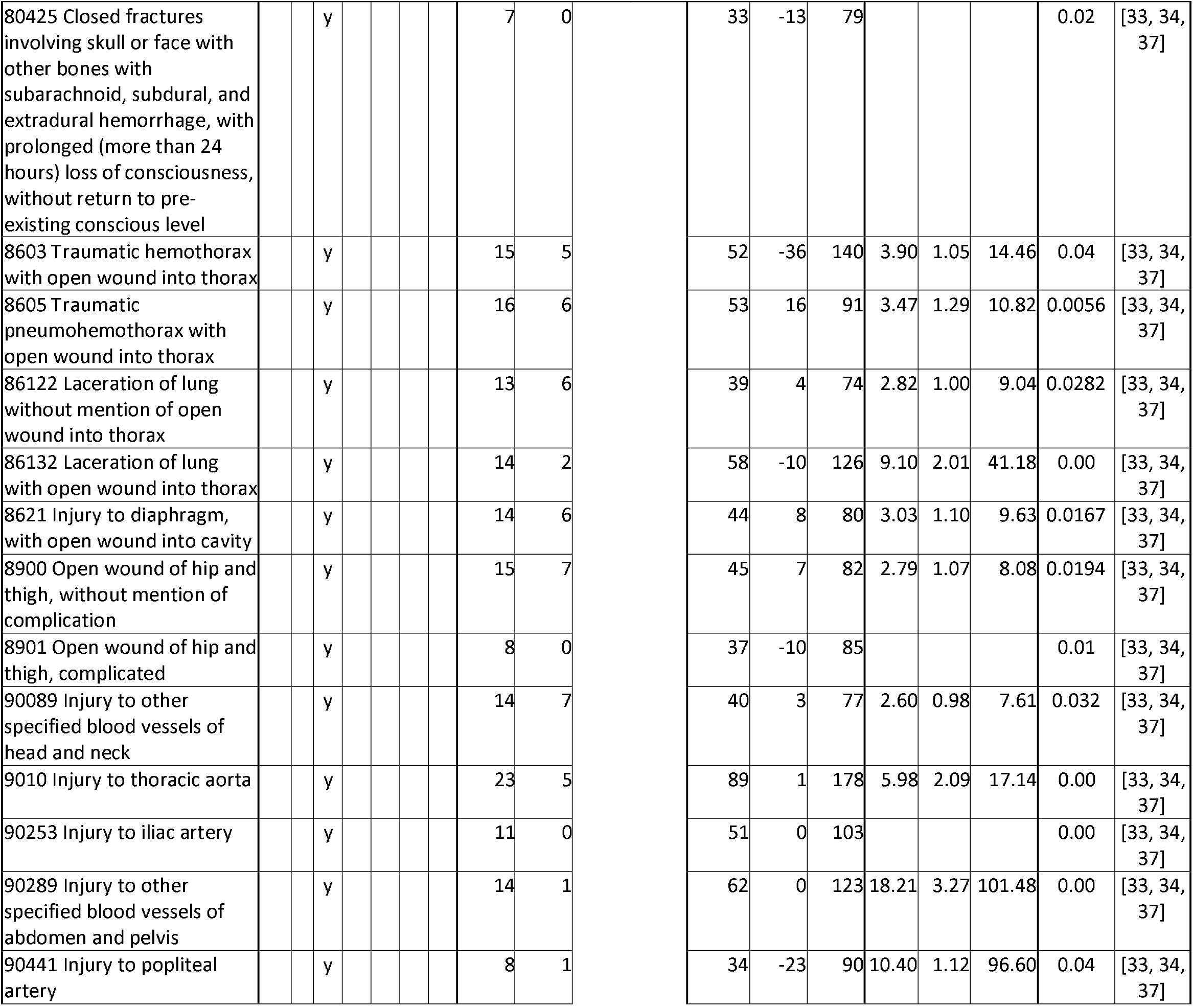
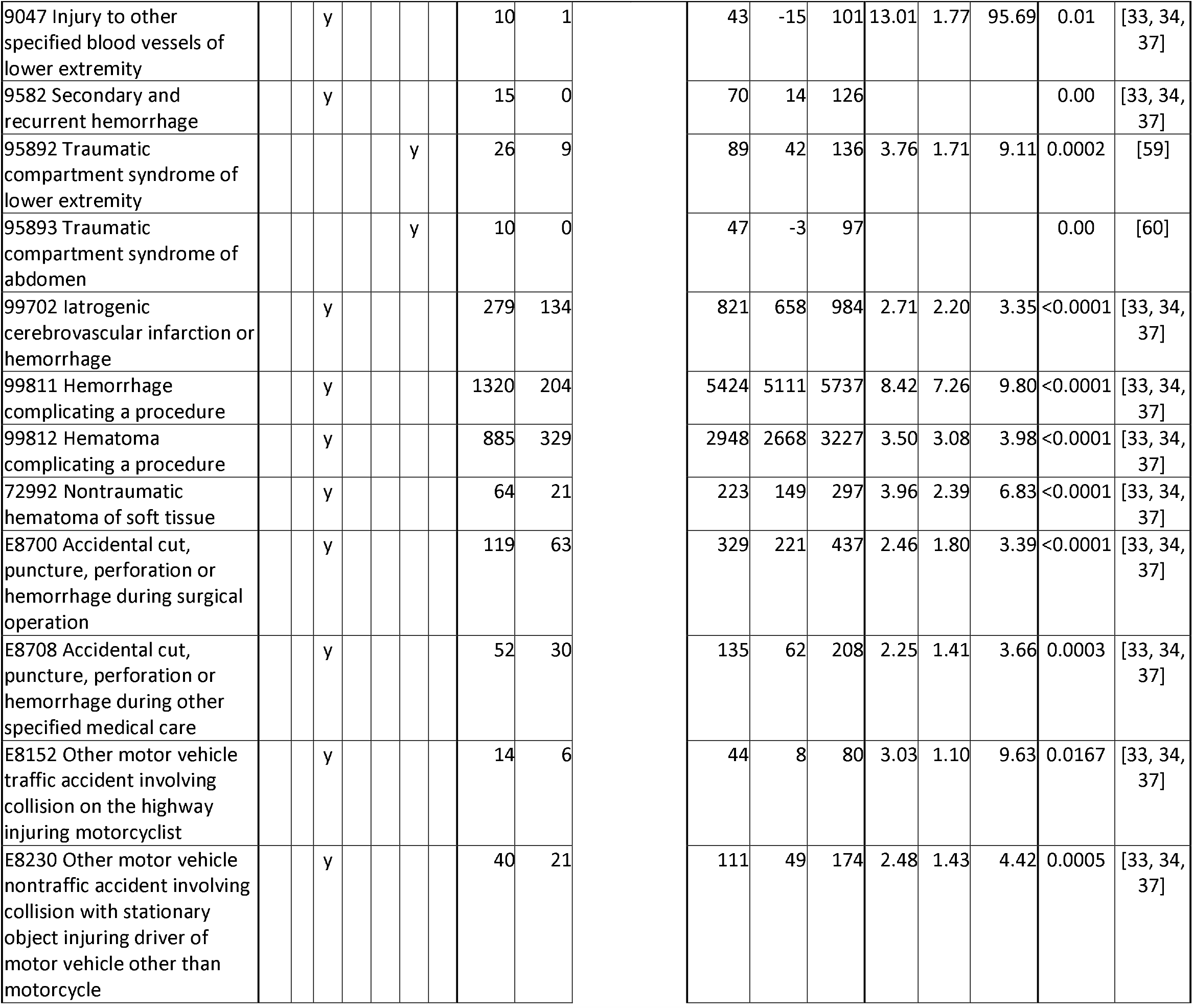
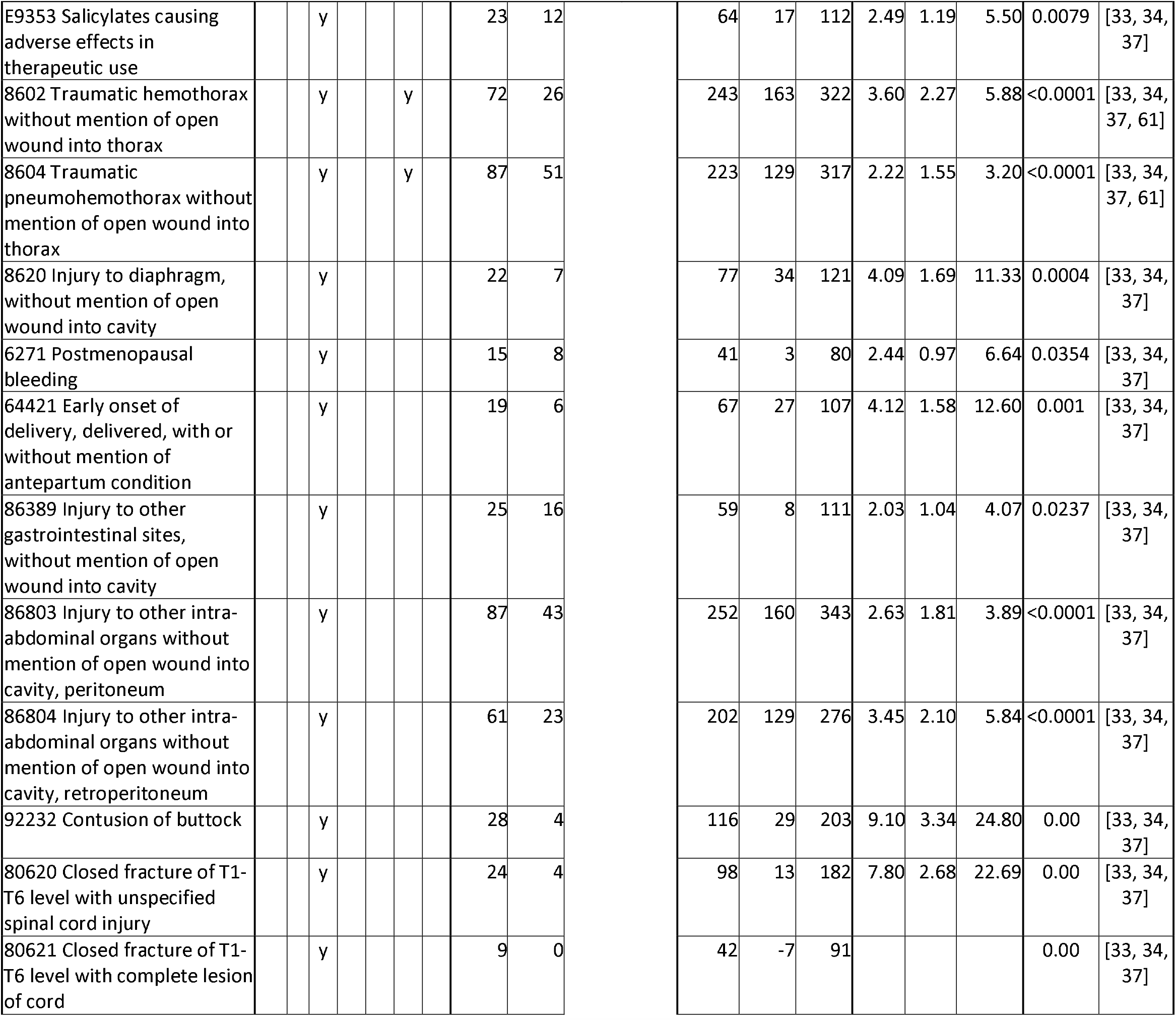
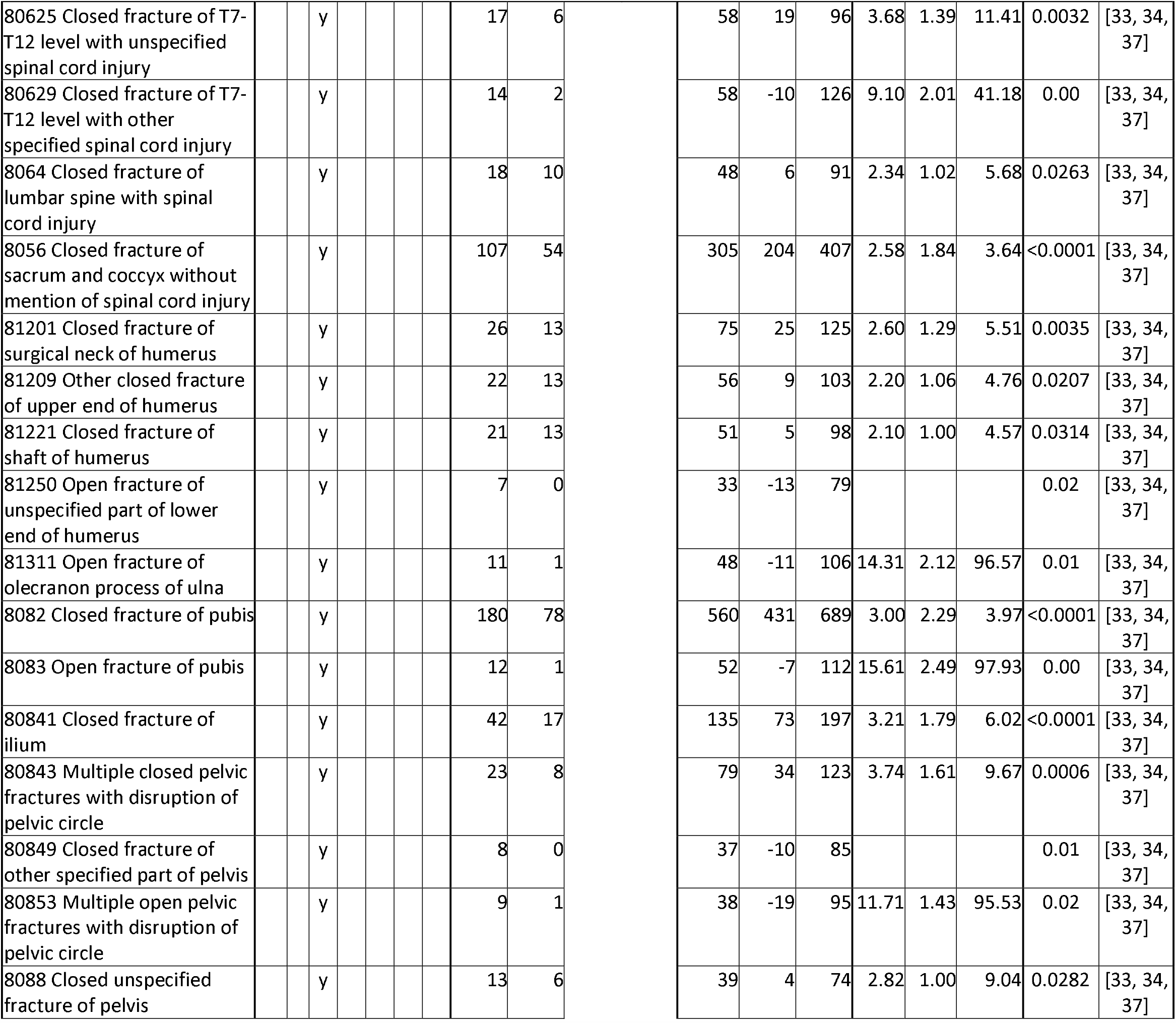
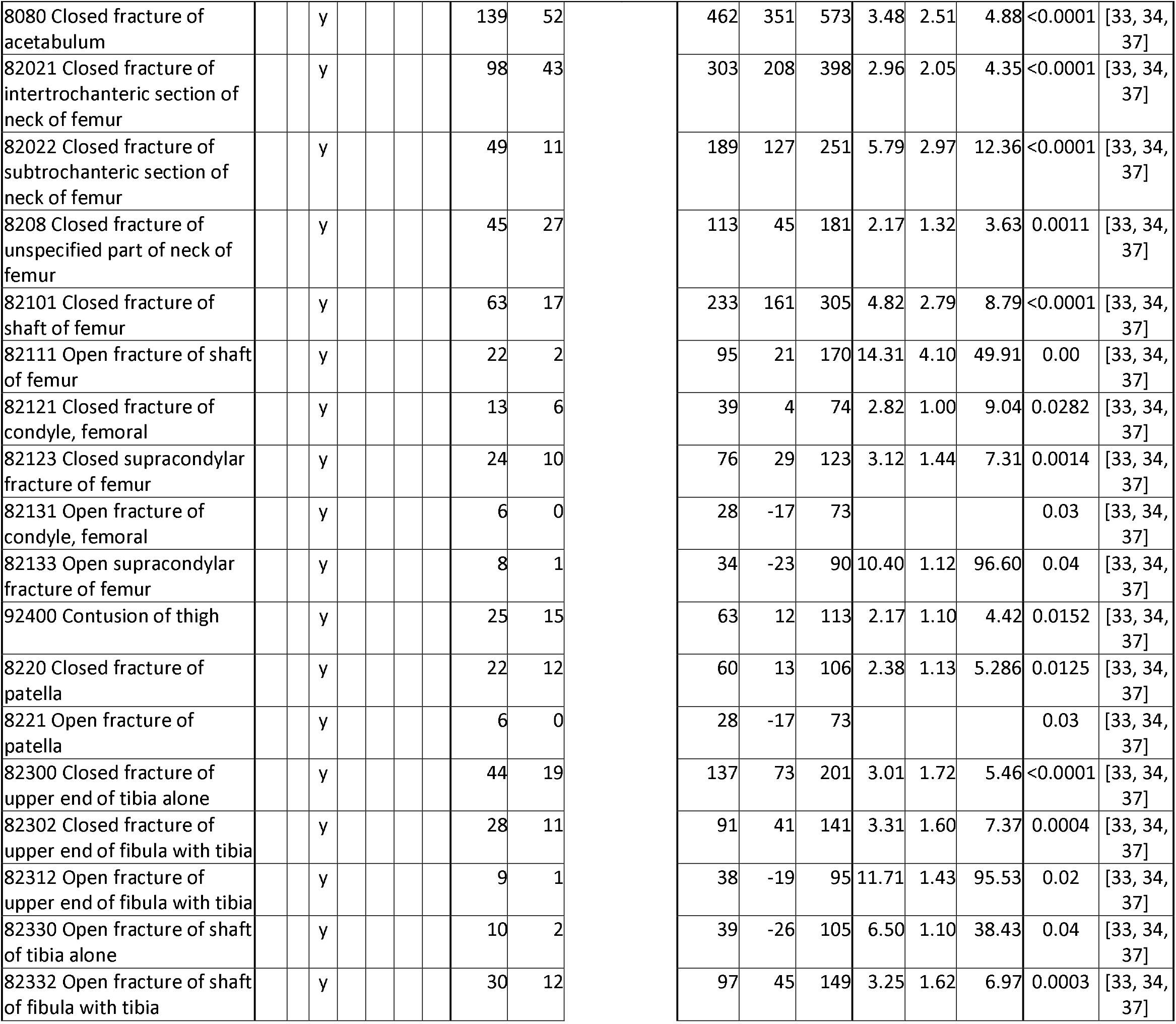
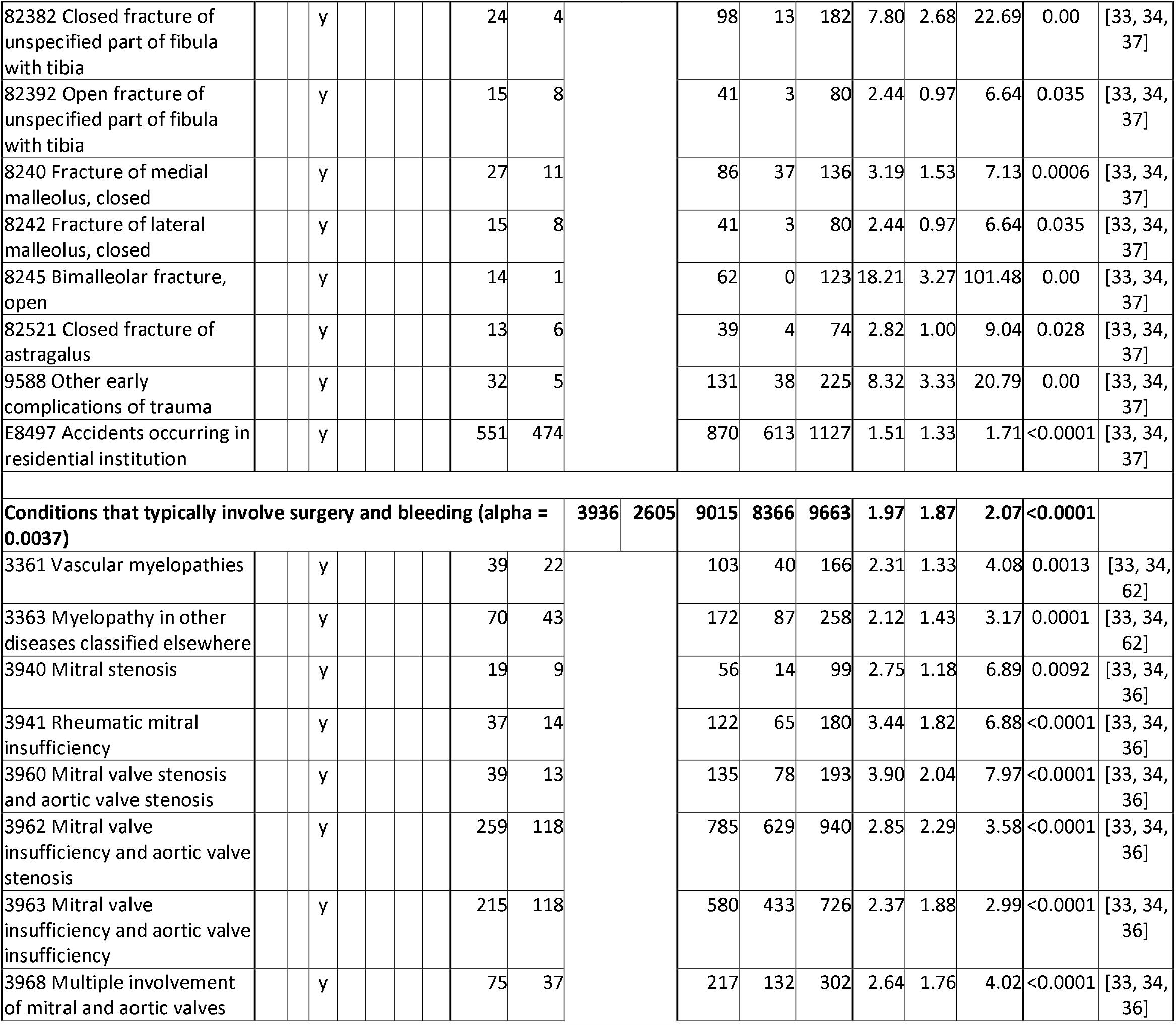
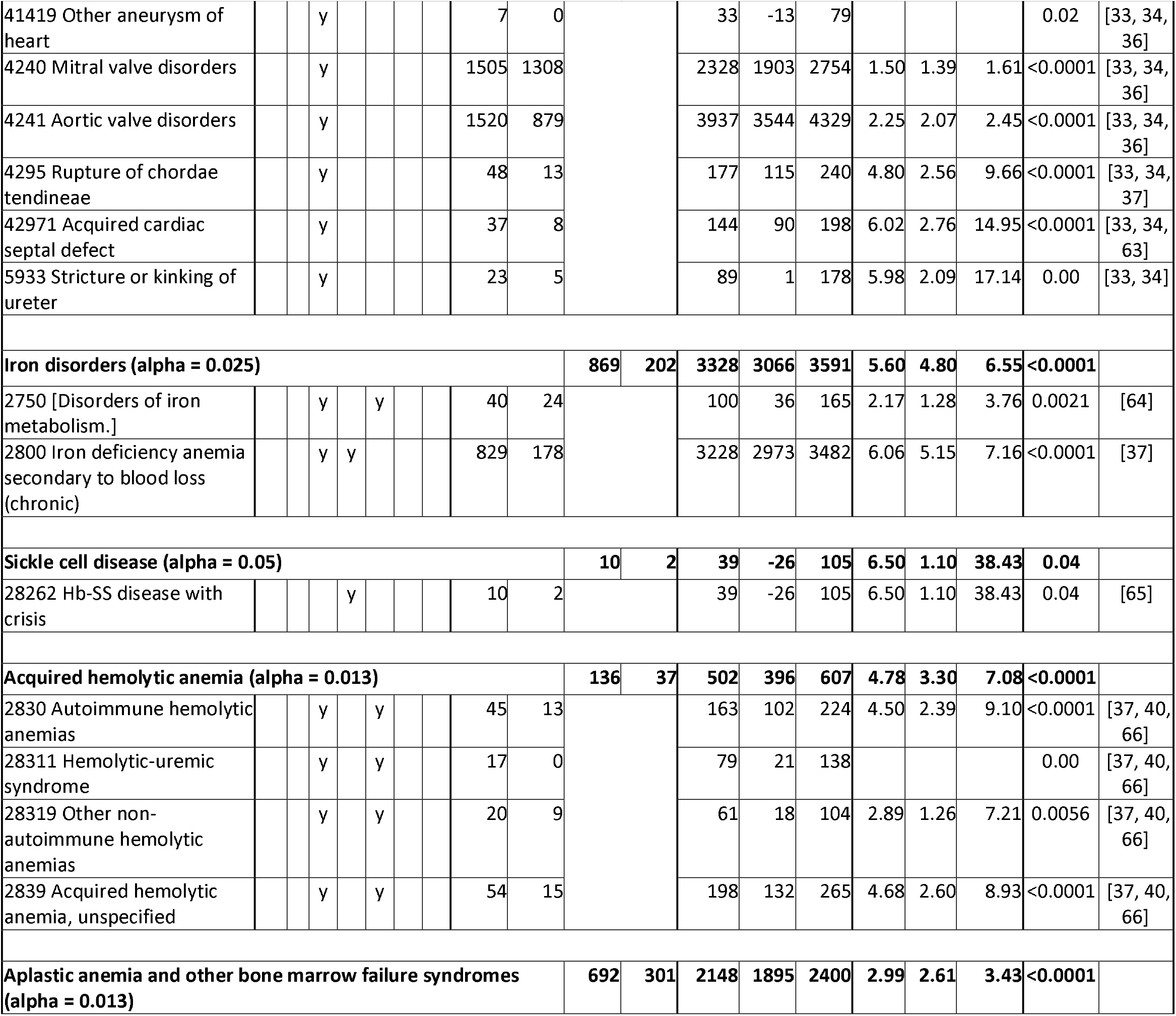
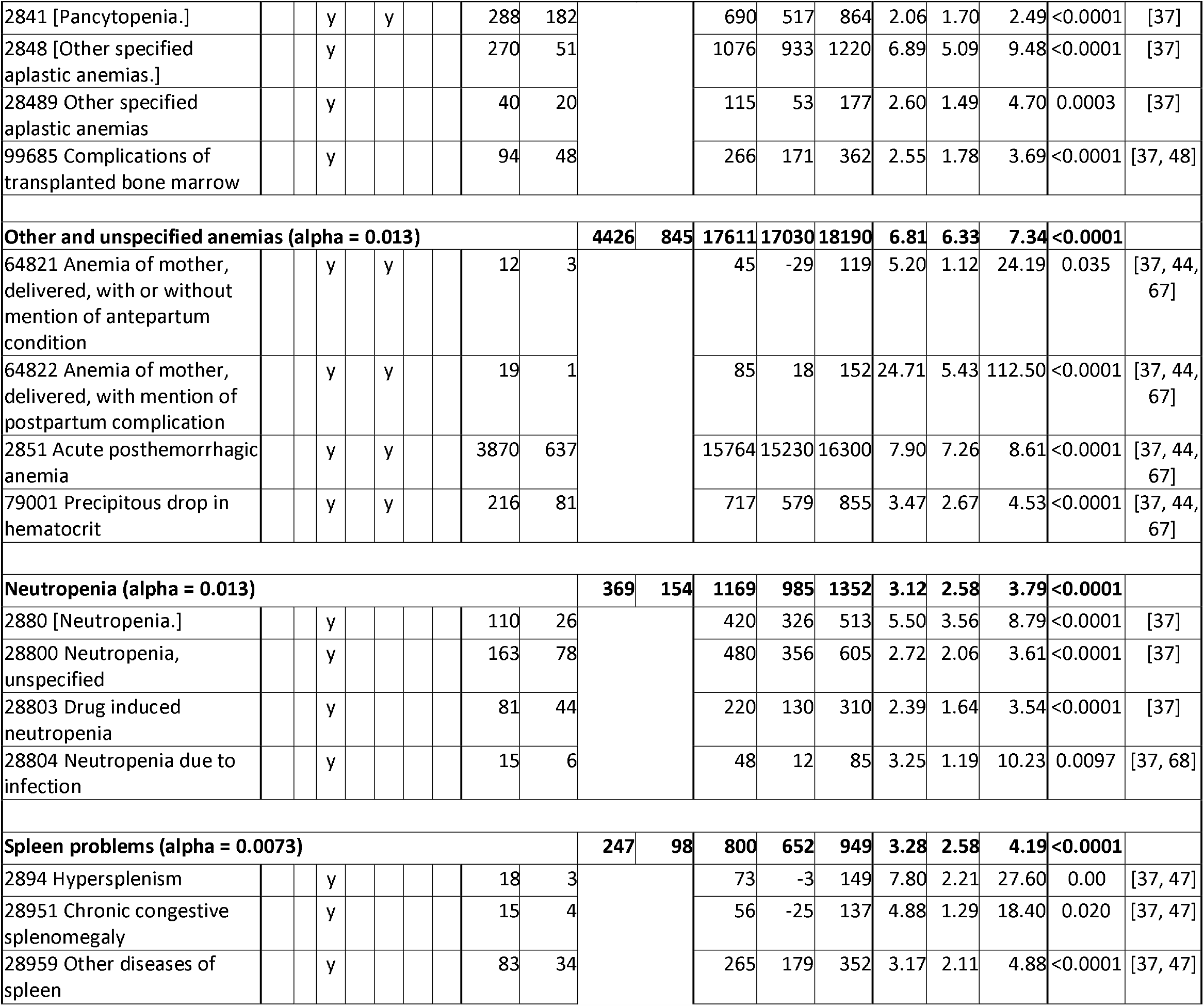
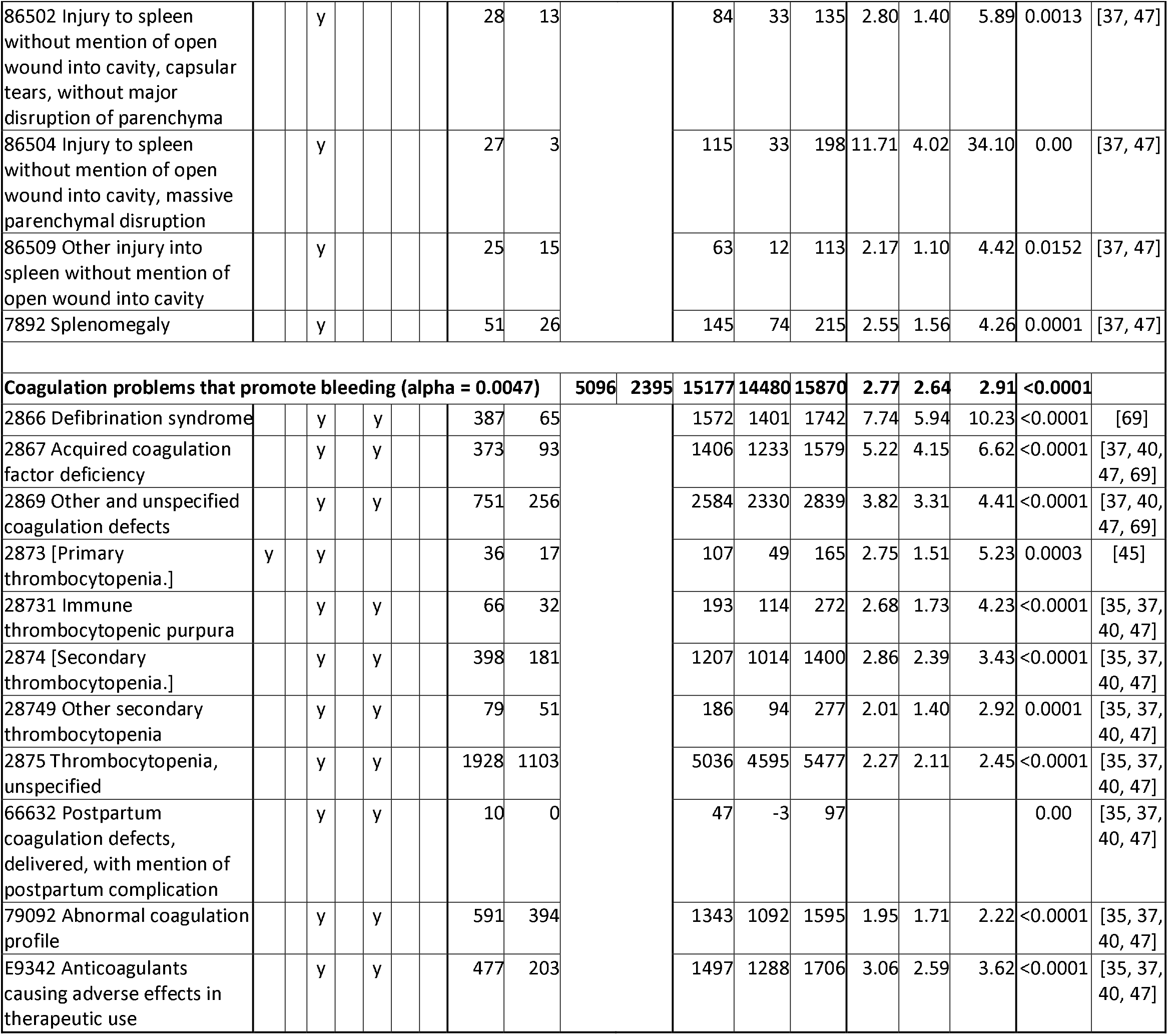
Analysis of T (transfusion) and C (comparison) groups for bleeding and anemia diagnosis codes. If all diagnoses in the table are dependent, alpha is 0.00026. Bracketed diagnosis code names were not in the MIMIC-III database, therefore were looked up [30] and confirmed in some notes.

**Table 5.**
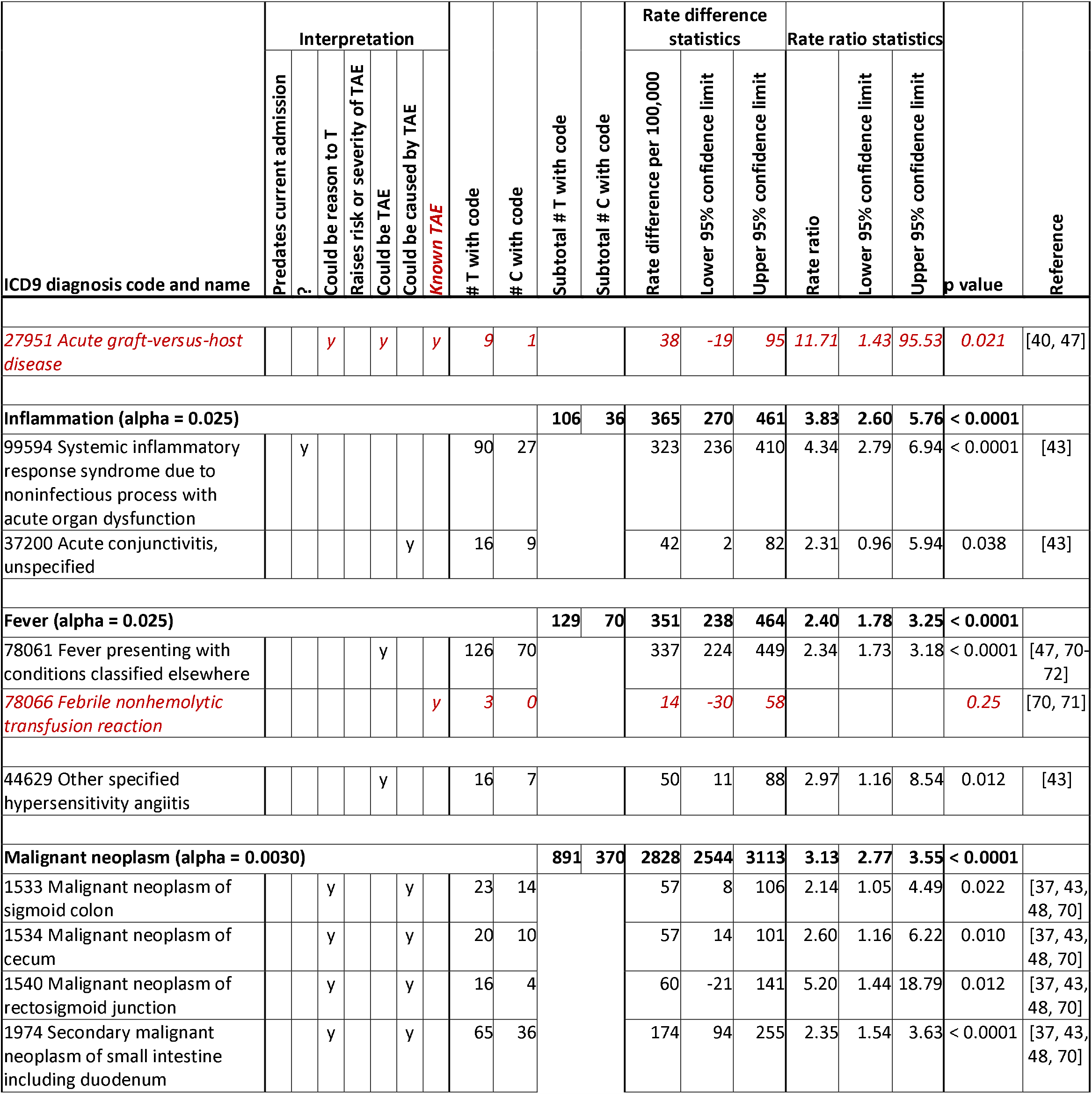
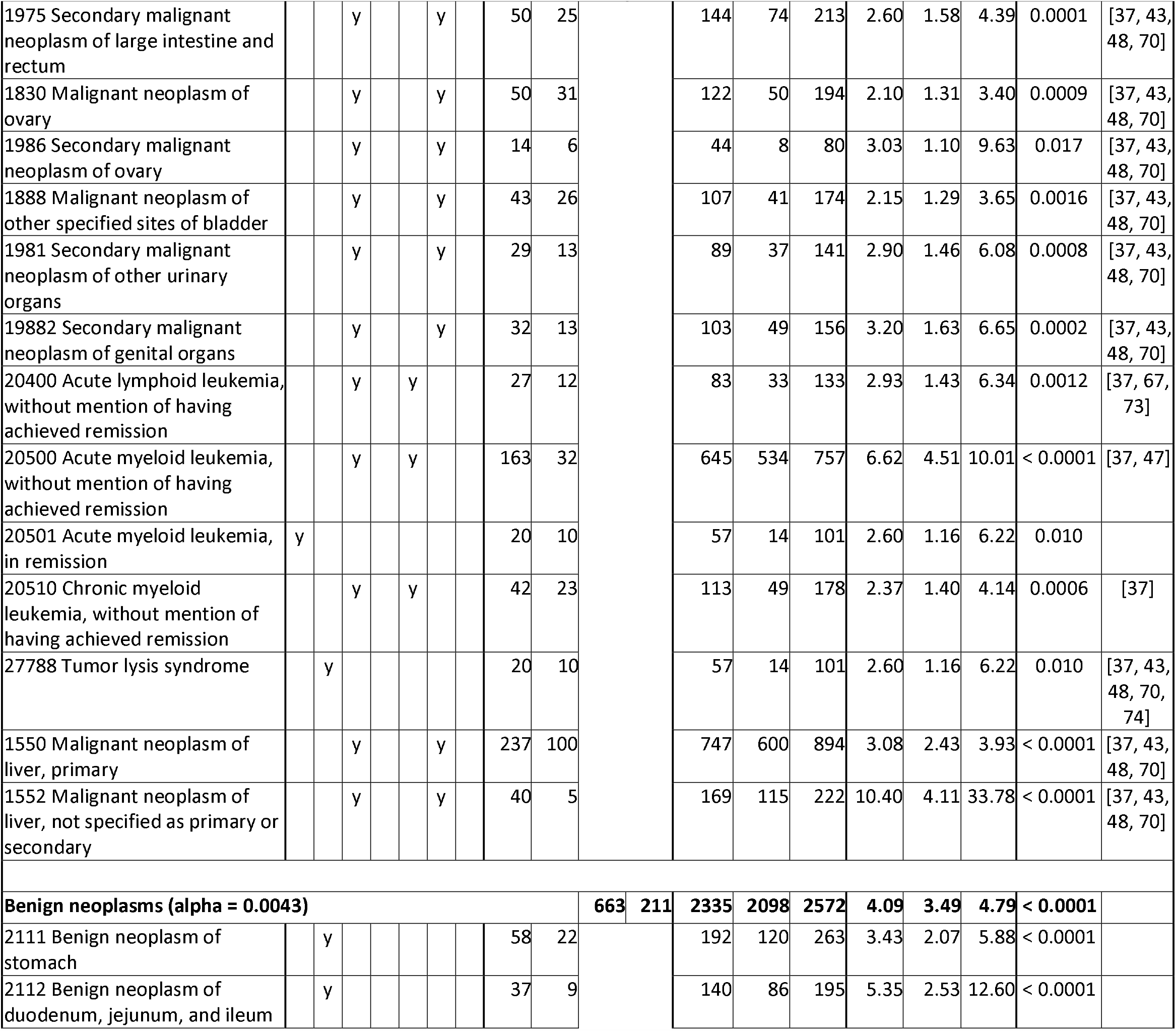
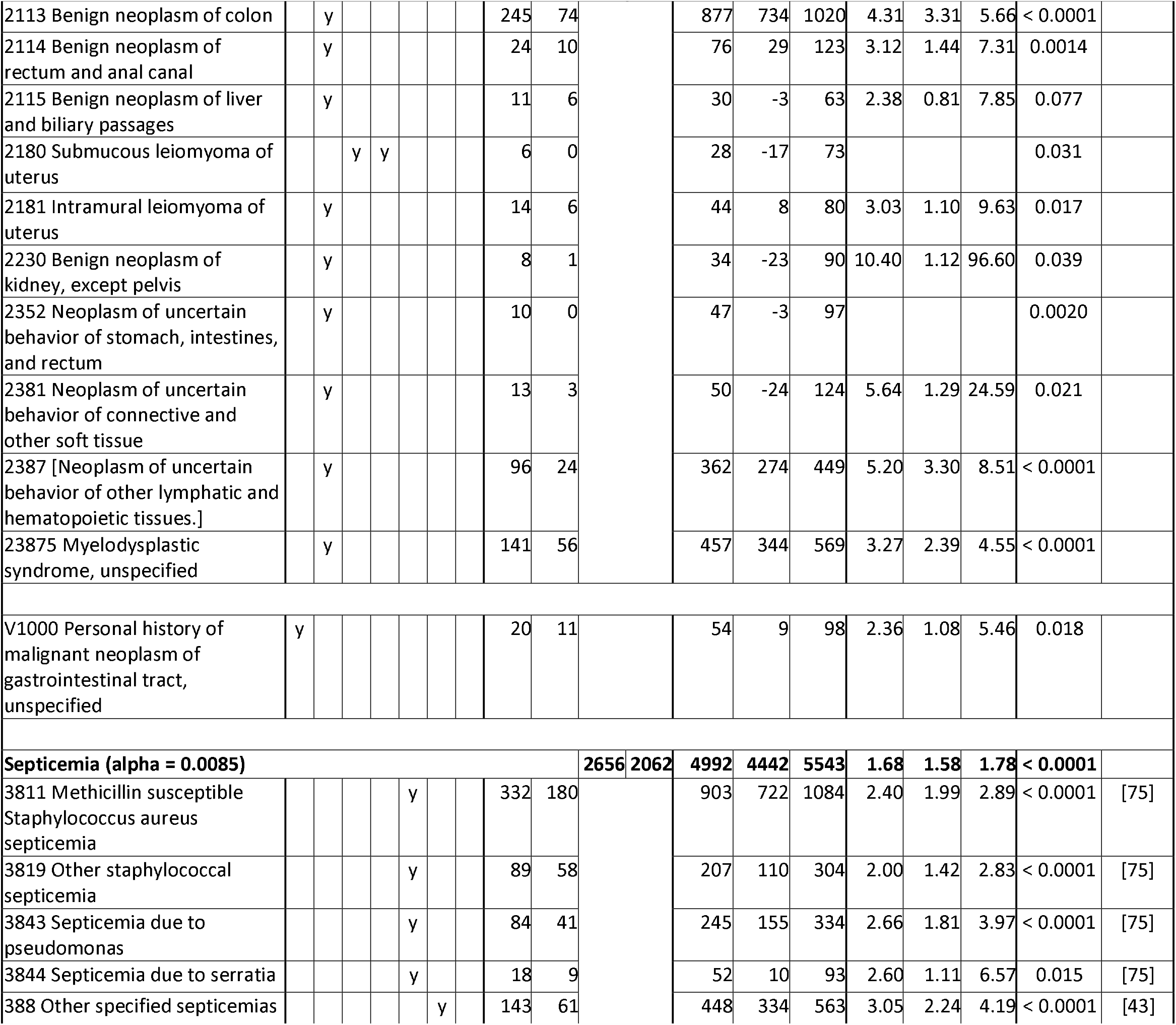
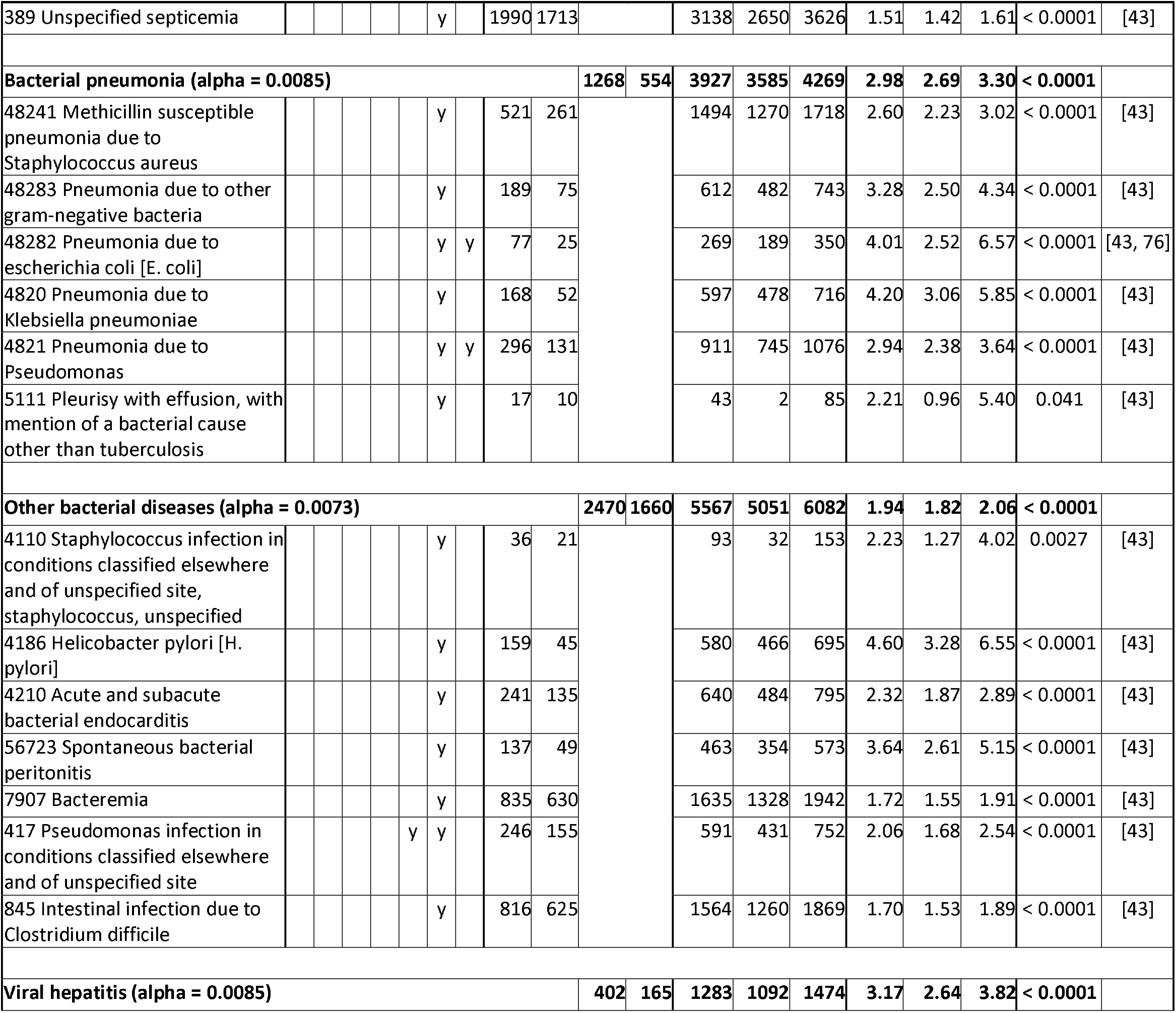
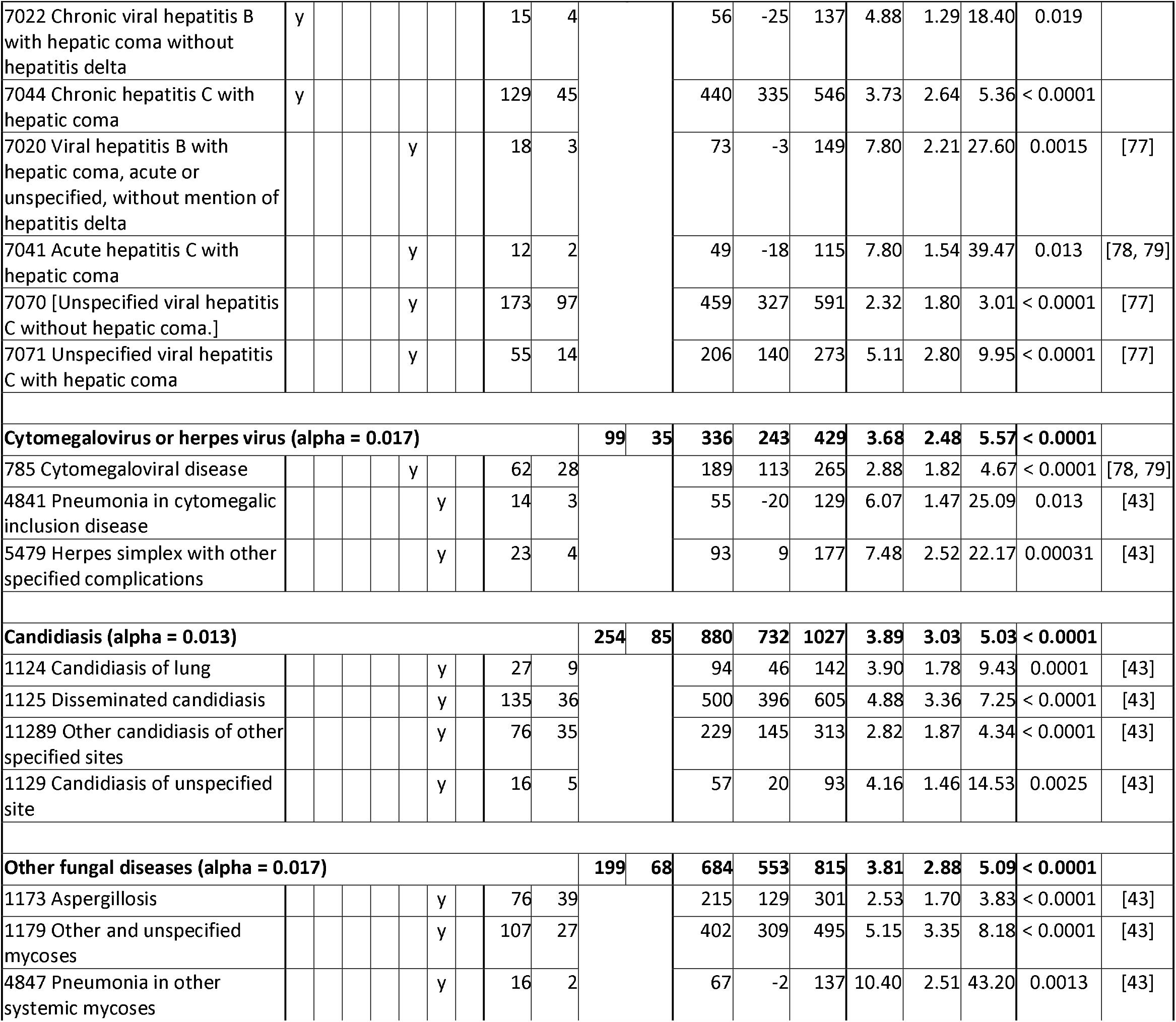
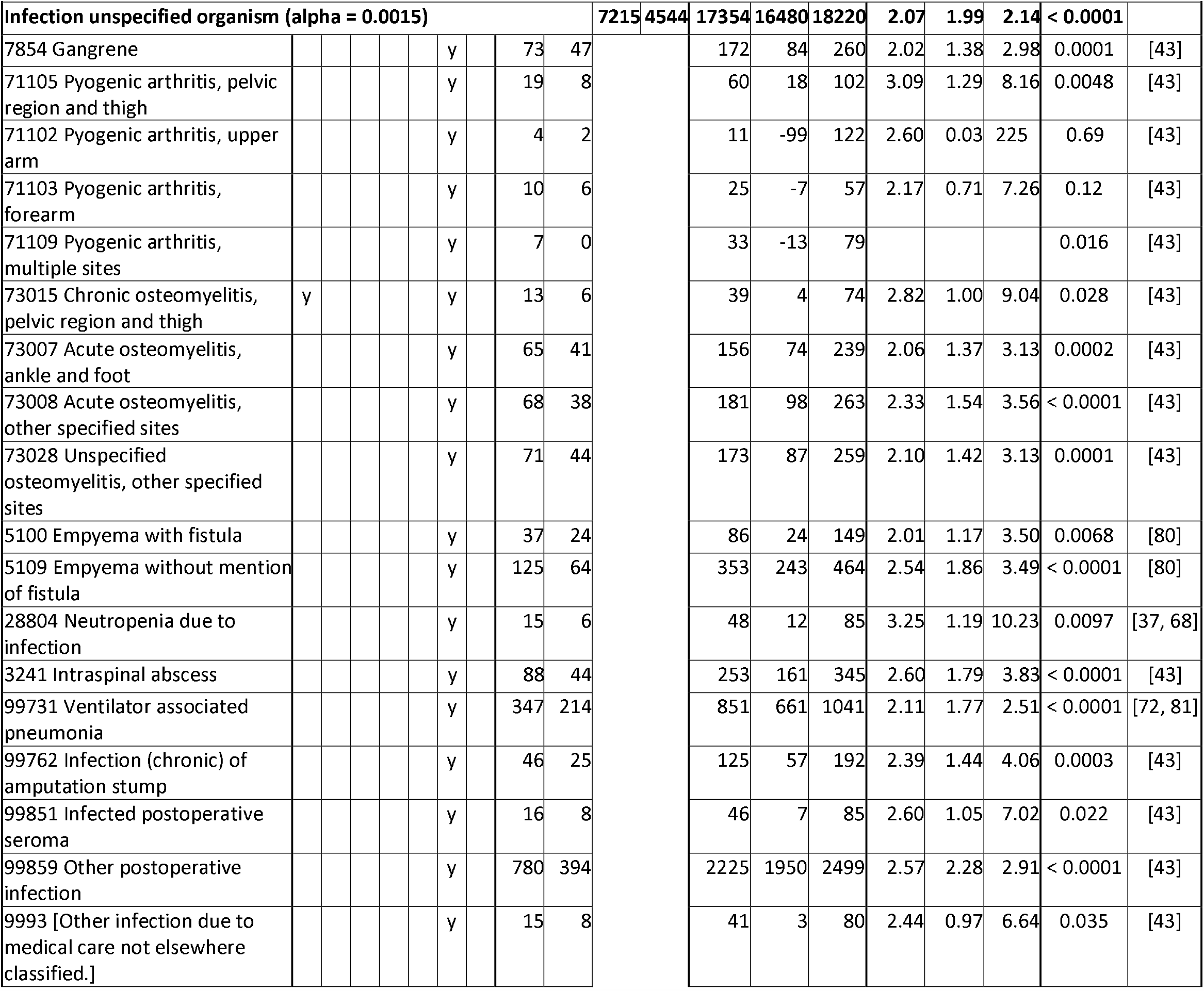
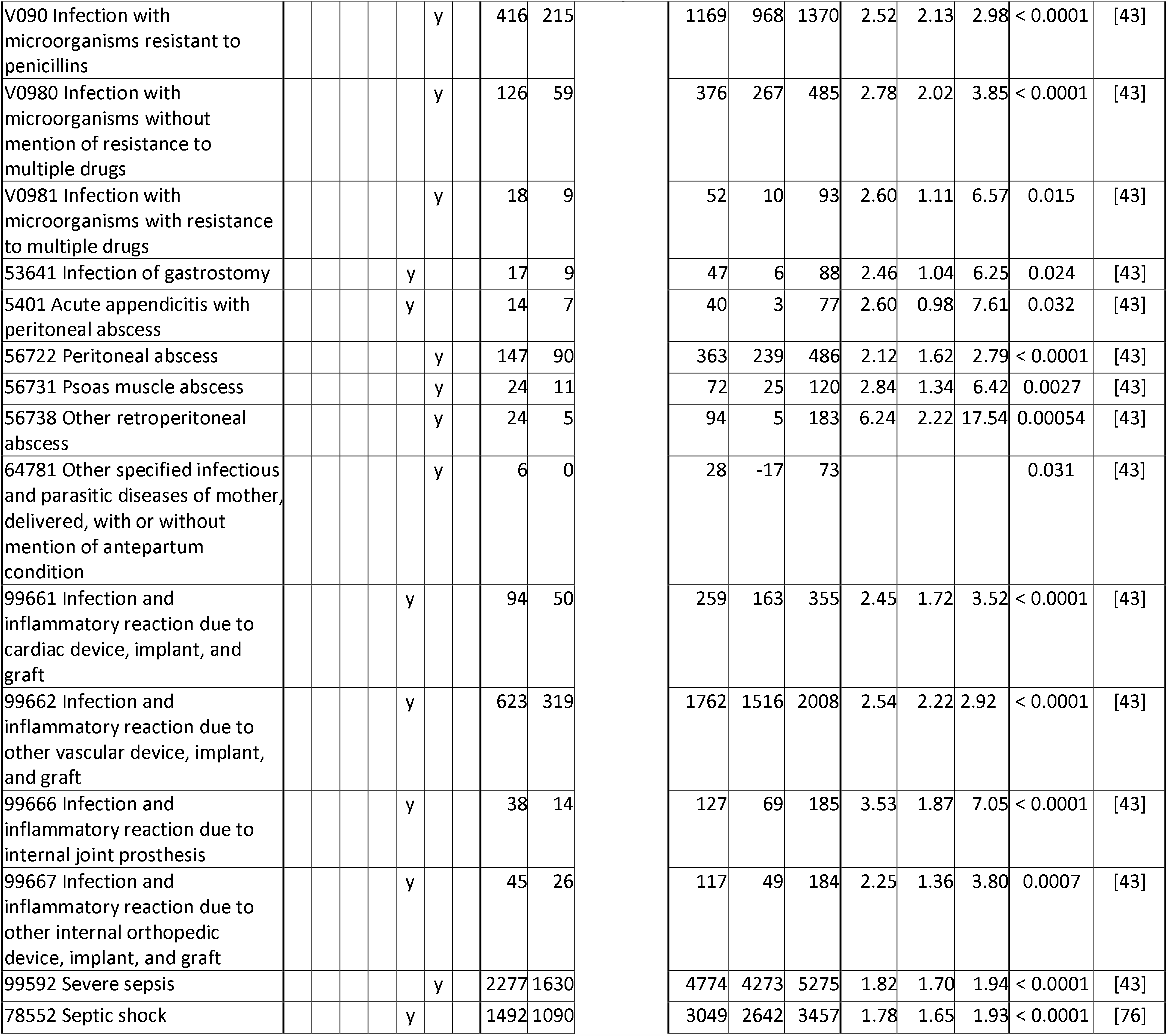
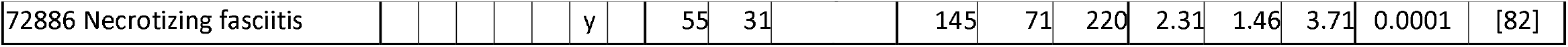
Analysis of T (transfusion) and C (comparison) groups for immune, neoplasm, and infection diagnosis codes. If all diagnoses in the table are dependent, alpha is 0.00049. Bracketed diagnosis code names were not in the MIMIC-III database, therefore were looked up [30] and confirmed in some notes.

**Table 6.**
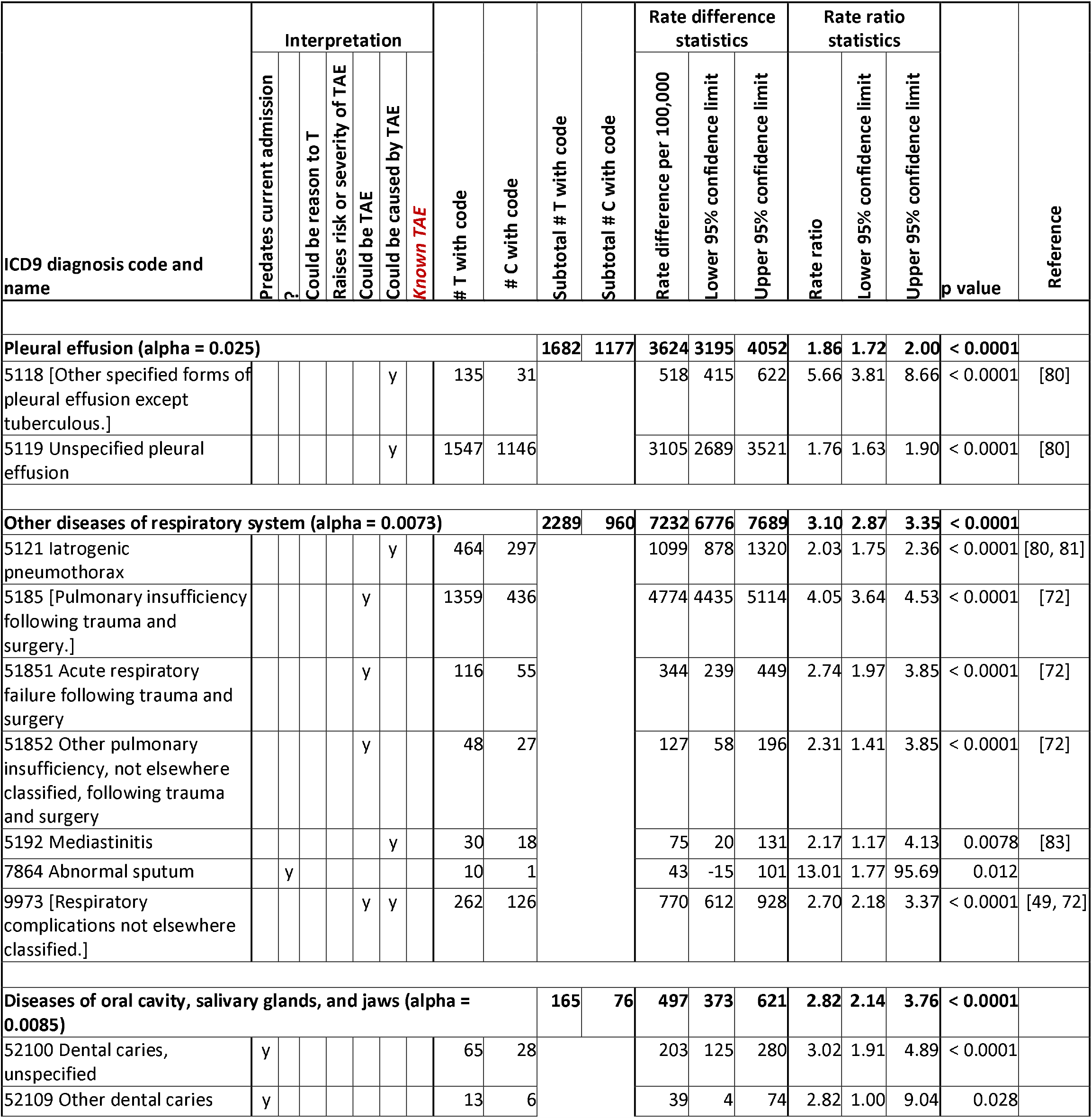
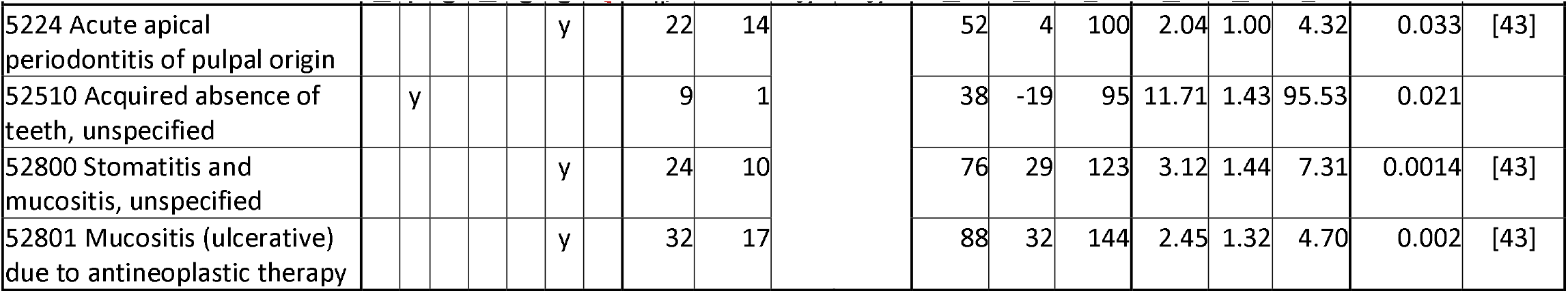
Analysis of T (transfusion) and C (comparison) groups for lungs and oral diagnosis codes. If all diagnoses in the table are dependent, alpha is 0.0034. Bracketed diagnosis code names were not in the MIMIC-III database, therefore were looked up [30] and confirmed in some notes.

**Table 7.**
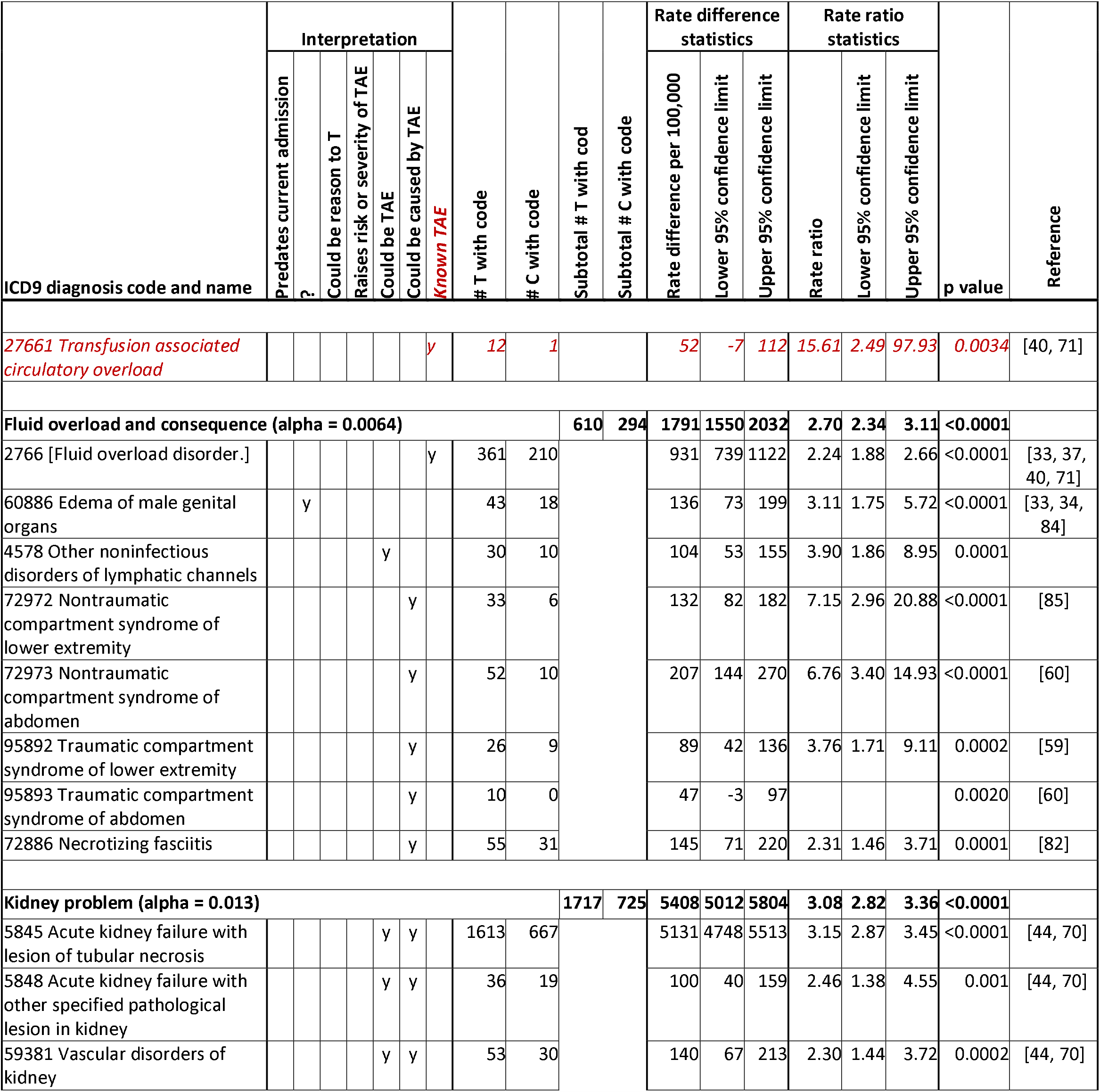
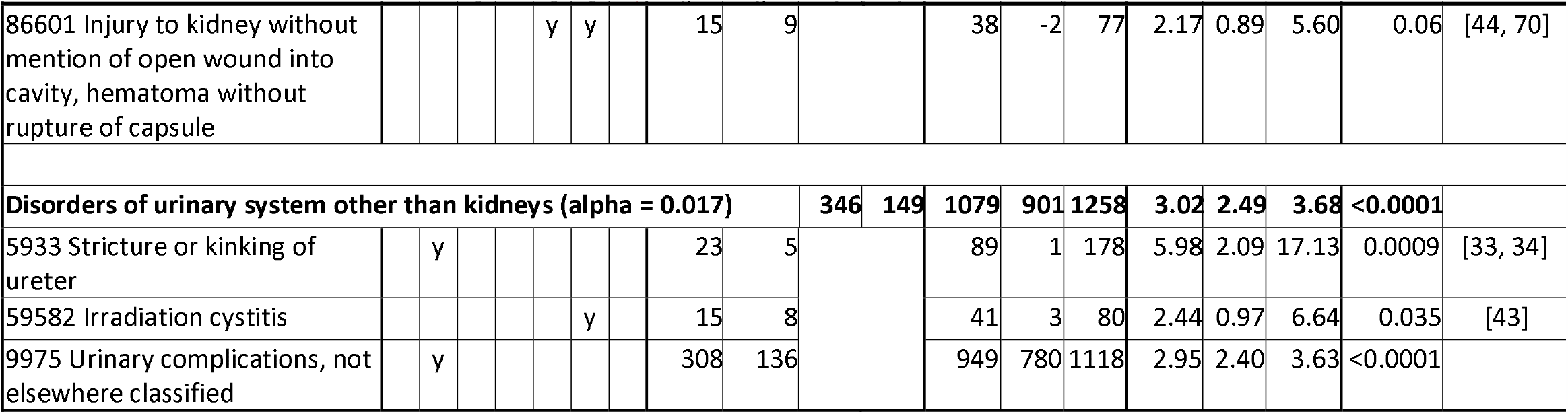
Analysis of T (transfusion) and C (comparison) groups for diagnosis codes that relate to fluids regulation. If all diagnoses in the table are dependent, alpha is 0.0032. Bracketed diagnosis code names were not in the MIMIC-III database, therefore were looked up [30] and confirmed in some notes.

**Table 8.**
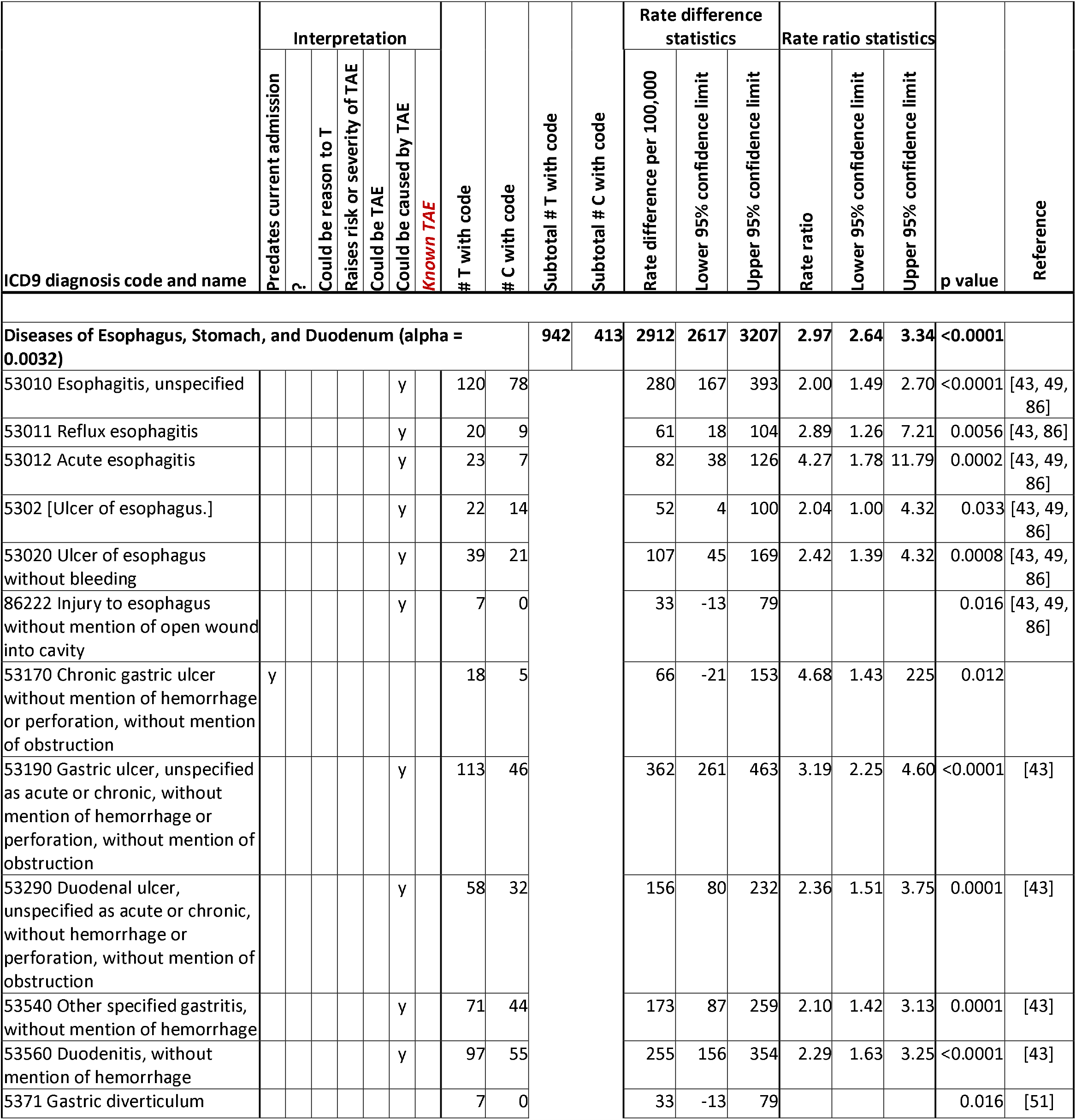
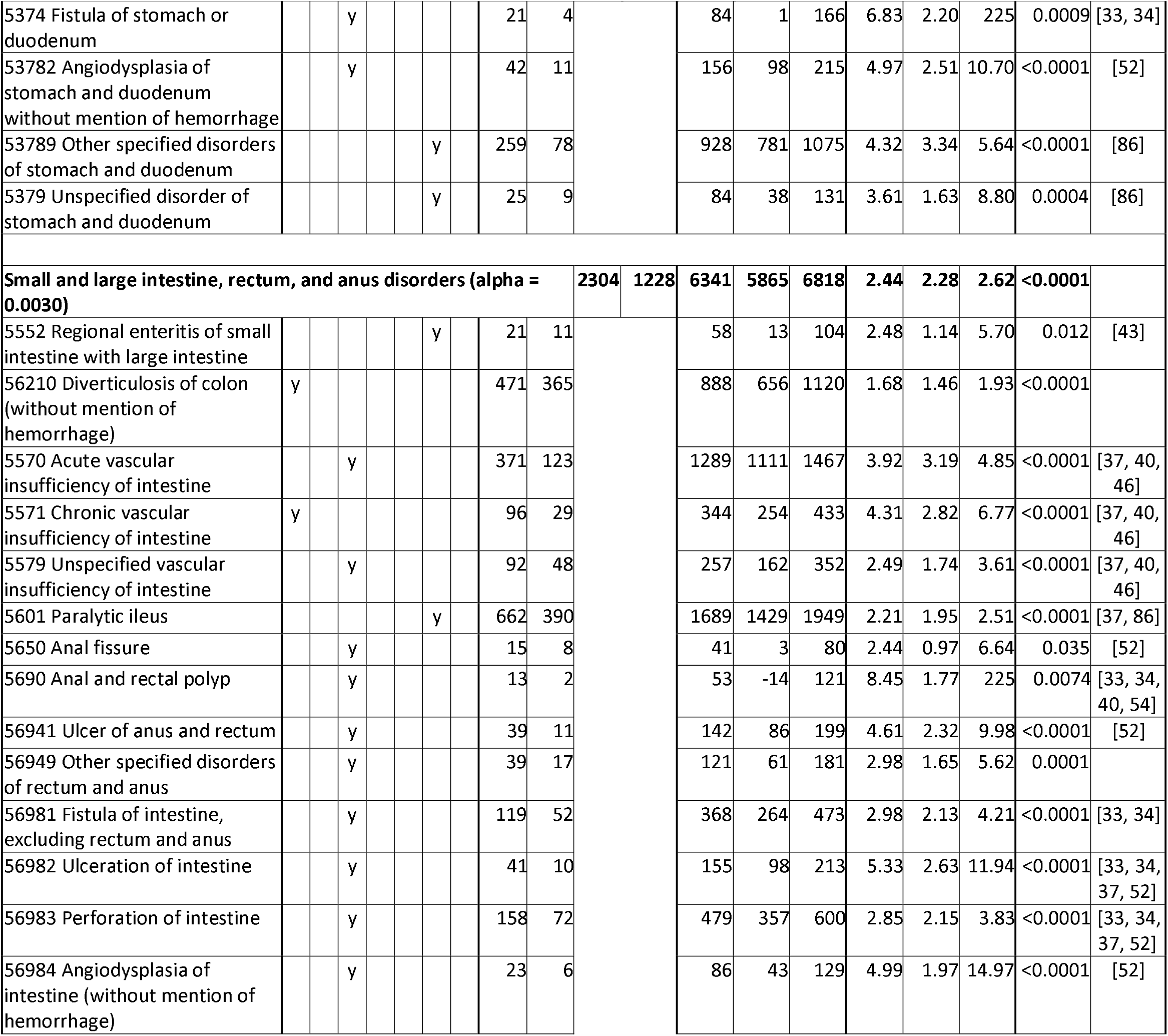
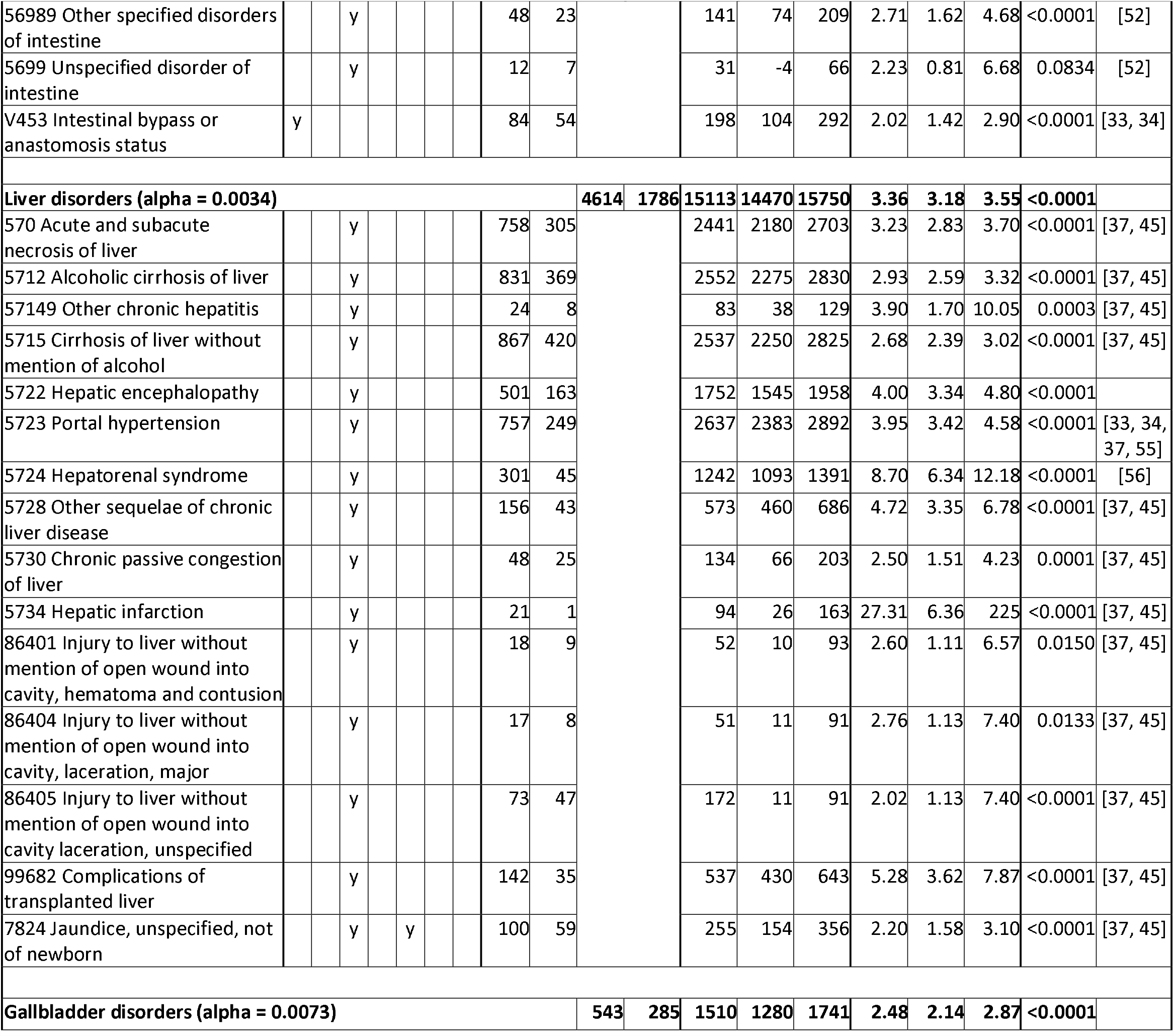
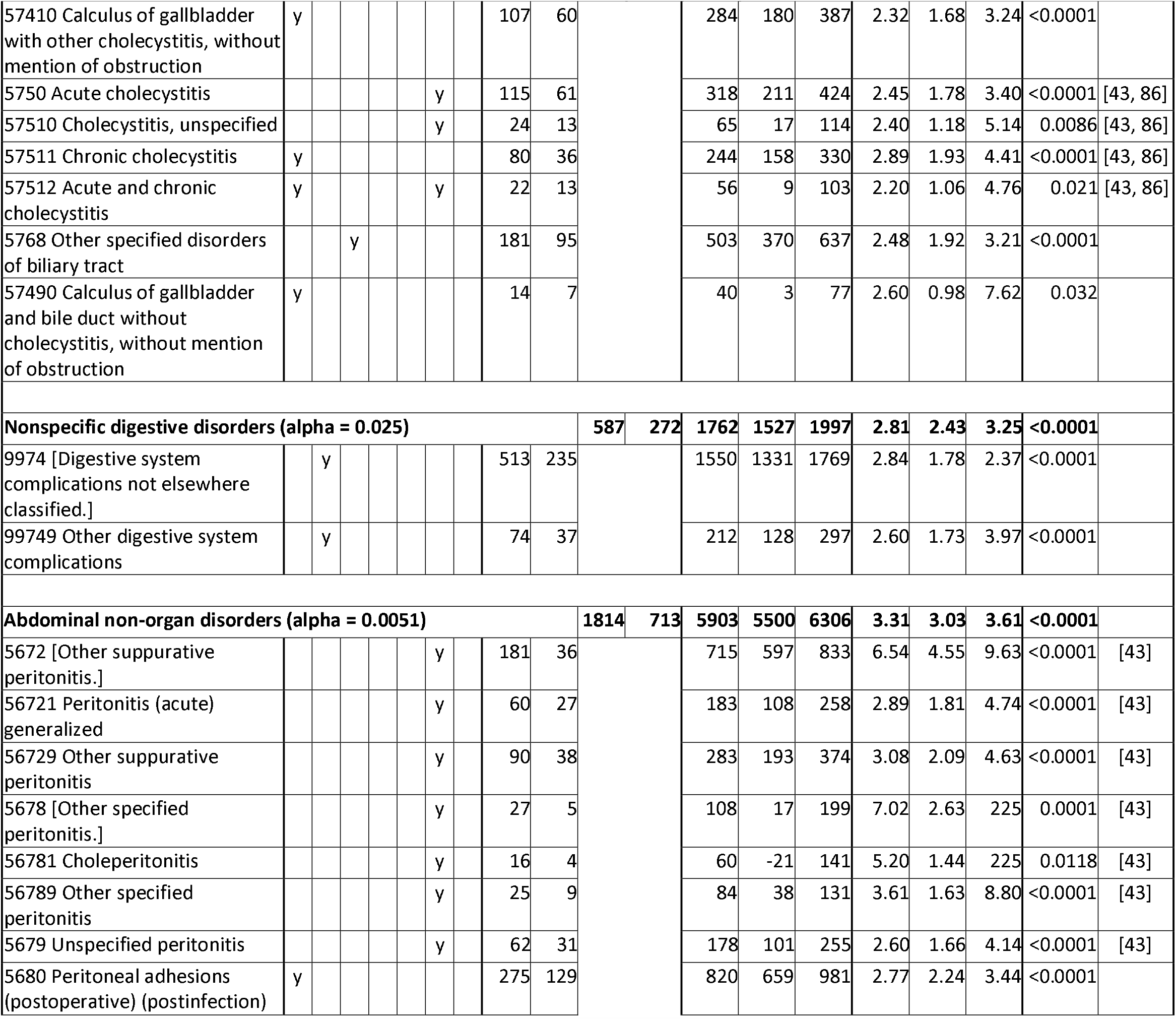
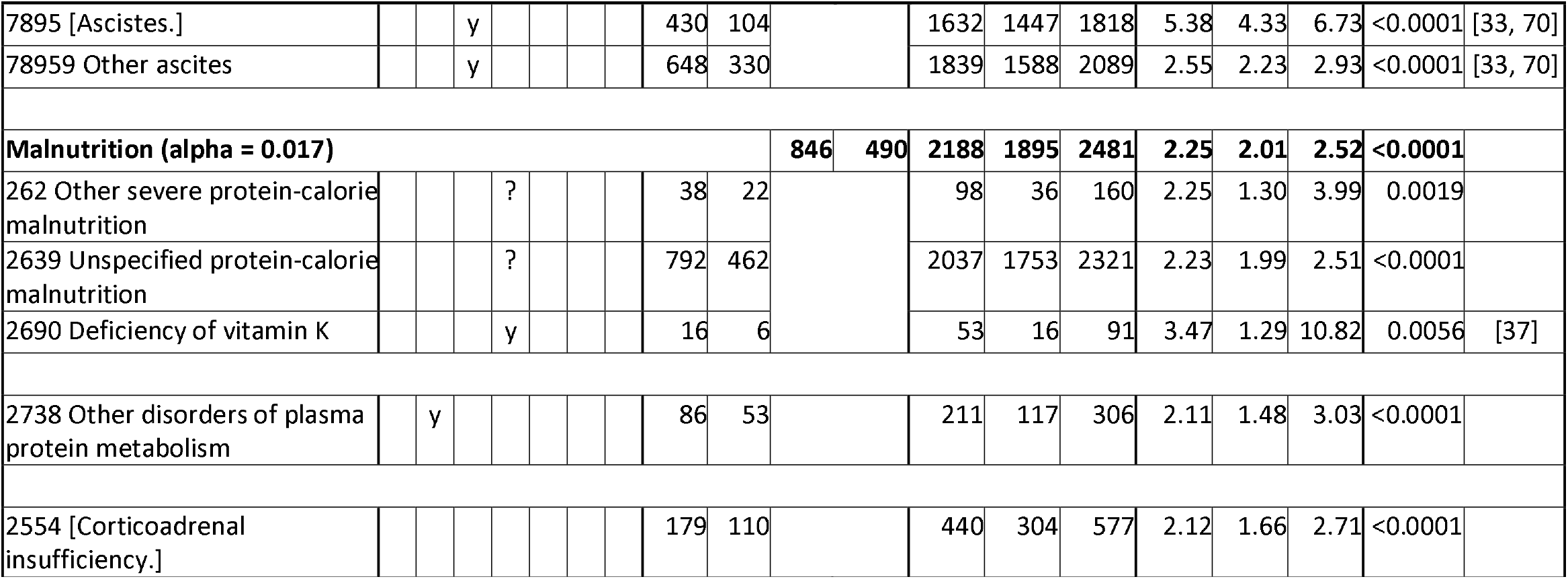
Analysis of T (transfusion) and C (comparison) groups for digestion and hormones diagnosis codes. If all diagnoses in the table are dependent, alpha is 0.00071. Bracketed diagnosis code names were not in the MIMIC-III database, therefore were looked up [30] and confirmed in some notes.

**Table 9.**
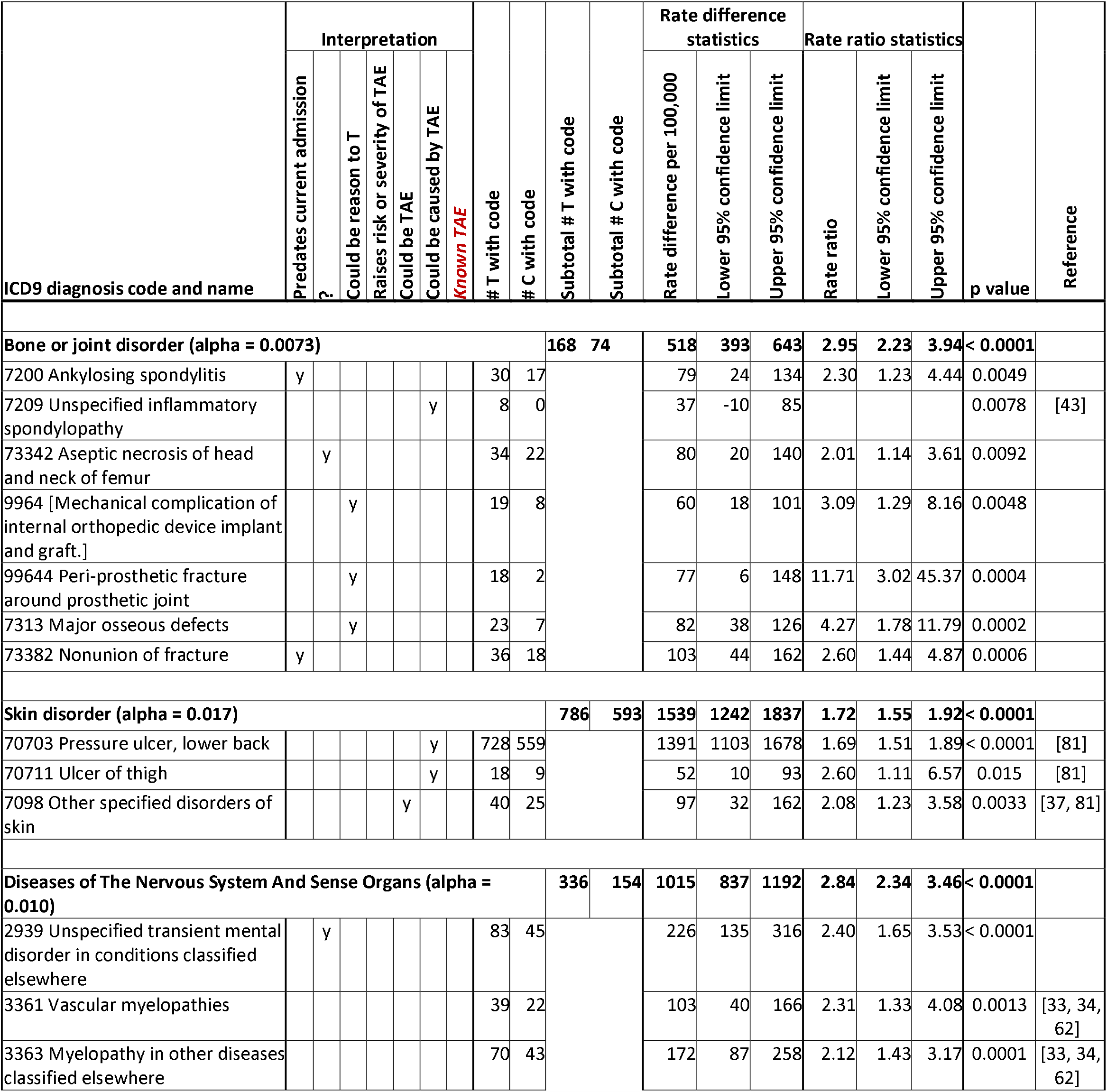
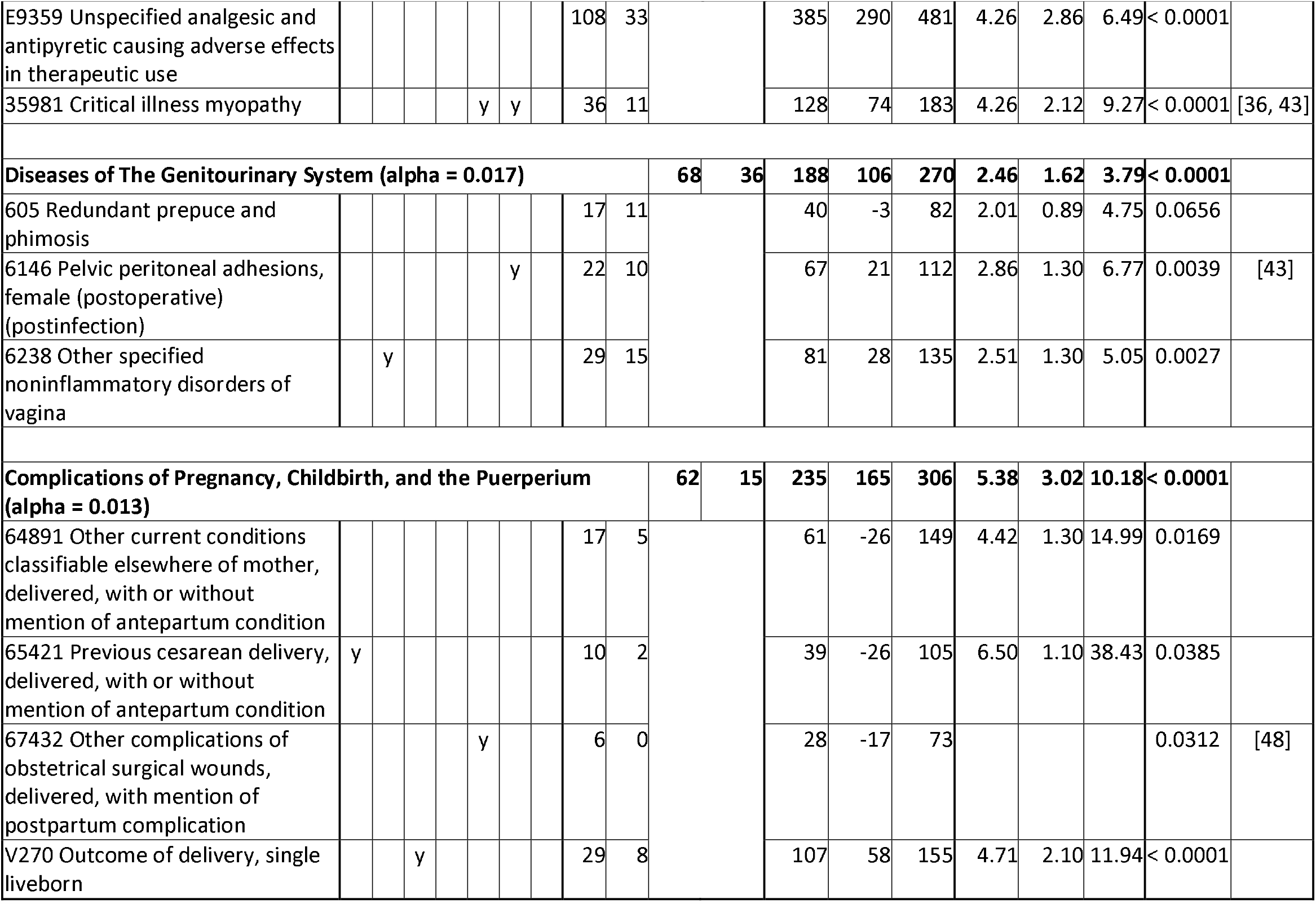
Analysis of T (transfusion) and C (comparison) groups for connective, nervous, and reproductive diagnosis codes. If all diagnoses in the table are dependent, alpha is 0.0023. TAE: transfusion adverse event. Bracketed diagnosis code names were not in the MIMIC-III database, therefore were looked up [30] and confirmed in some notes.

There were codes for conditions similar to TAE that were statistically significantly more common in T:

- Some of the hypercoagulation and emboli disorders in Table 3 and bleeding in Table 4 may have been alloimmune thrombocytopenic purpura [35].
- Some of the anemia diagnoses in Table 4 may have been hemolytic transfusion reactions [37, 40].
- Some of the infections in Table 5 may have been transmitted via transfusion 43, 75-79].
- Codes in Table 6 that might have signified TRALI [65] were:
  - 5119 (Unspecified pleural effusion) [80]
  - 5118 (Other specified forms of pleural effusion except tuberculous) [80]
  - 9973 (Respiratory complications not elsewhere classified) [49, 72]
  - If transfusion was associated with the trauma or surgery [72]
    ▪ 5185 (Pulmonary insufficiency following trauma and surgery)
    ▪ 51851 (Acute respiratory failure following trauma and surgery)
    ▪ 51852 (Other pulmonary insufficiency, not elsewhere classified, following trauma and surgery)

Many other codes may have been consequences of TAE, for example:

- Kidney injury [44, 70] in Table 7, such as:
  - 5845 (Acute kidney failure with lesion of tubular necrosis)
  - 5848 (Acute kidney failure with other specified pathological lesion in kidney)
- Esophagus injury or gastritis in Table 8 possibly due to intubation, such as:
  - 53010 (Esophagitis, unspecified) [43, 49, 86]
  - 53540 (Other specified gastritis, without mention of hemorrhage) [43]
- Pressure ulcer [81] due to extended bed rest in Table 9:
  - 70703 (Pressure ulcer, lower back)
  - 70711 (Ulcer of thigh)

As one would expect, many of the diagnosis codes that were statistically significantly more common in T than C were, or could have been indicators of, reasons to transfuse (including preexisting conditions:

- Loss of blood due to spontaneous reasons, trauma, or surgery [33-35, 37, 47, 48], in Table 4
- Malignant cancers [37, 43, 47, 48, 70] in Table 5

Some codes might have been preexisting conditions that raise the risk or severity of TR, for example:

- 28262 (Hb-SS disease with crisis) [65] in Table 4
- 2690 (Deficiency of vitamin K) [37] in Table 8

Some codes may represent both reasons to transfuse and consequences of transfusion, for example:

- Emboli and thrombi in Table 3 [76]
- Bleeding in Table 4 [35, 37, 47, 48]
- Hemolytic anemia in Table 4 [37, 76]

There are many codes that we were not able to explain. Most were vague, while some were specific, for example:

- Benign neoplasms in Table 5
- 5933 (Stricture or kinking of ureter) in Table 7
- 73342 (Aseptic necrosis of head and neck of femur) in Table 9.

Many medical care AEs occur at higher frequency in hospital critical care [91, 92].

As would be expected, many diagnosis codes that were associated with transfusion were reasons to transfuse or predated the admission.

A relatively small number were explicit TAE codes. Some codes were for conditions that are the same as explicit TAE codes, but without the attribution and many of them may have been TAE. The strategy of looking for codes for conditions similar to each other has been used to analyze adverse event reports [93]. One interpretation of this phenomenon is knowledge that it is TAE but reluctance to code as such; another could be lack of awareness that the condition was a TAE, and a remaining possibility is that the potential TAE occurred before transfusion.

Our findings on two TAE codes were statistically significantly less than would have been expected from Medicare studies of the frequencies of TRALI and febrile non-haemolytic transfusion reaction (ICD9 code 78066) [20, 90] (see Table 10). Our results were not statistically significantly different for TACO and posttransfusion purpura (ICD9 code 28741) [19, 89].

**Table 10.** Comparison of MIMIC-III diagnoses of TAEs (transfusion adverse events) with published Medicare data from varying time periods 2007-2012. The expected number applies the rate found in Medicare data to the study T (transfusion) group.

| Transfusion adverse event (TAE) | # TAE in T | Medicare data |  |  | Expected # in T | Fisher's exact test (FET) p value |
| --- | --- | --- | --- | --- | --- | --- |
|  |  | # TAE/<br># transfused | Calendar years | Reference |  |  |
| Transfusion related acute lung injury (TRALI) | 0 | 2556/<br>11,378,264 | 2007-2011 | [20] | 4.8 | 0.019 |
| Transfusion associated circulatory overload (TACO) | 12 | 1340/<br>2,147,038 | 2011 | [19] | 13.4 | 0.089 |
| Posttransfusion purpura | 0 | 78/<br>4,336,338 | 2011-2012 | [89] | 0.39 | 1 |
| Febrile non-haemolytic transfusion reaction | 3 | 2517/<br>4,336,338 | 2011-2012 | [90] | 12.5 | 0.004 |

Our results showed that allowing additional diagnoses that could indicate TAE could be useful for widening the net for detection of signals of TAEs.

Some diagnoses may have been consequent to TAEs.

A number of diagnoses were not explained.

Our investigation of TAE codes in C demonstrated that codes alone may not provide the full clinical picture that is in the notes. Other studies have reported that while codes can provide more information than AE reports [4], the codes themselves provide an incomplete picture [12]. Our main measures of RD and RR could be underestimates of the risks of T if the loss of information due to reliance on codes is evenly distributed across the data or otherwise biased.

The analysis of codes is merely to generate signals of AE. All signals require further investigation to assess causality because diagnosis codes generally do not provide the sequence of events before, and never during, hospitalization or assess competing explanations of the associations.

## CONCLUSIONS

This study successfully demonstrated simple surveillance of patient harm by using diagnosis codes in a large healthcare database, as a supplement to current post-marketing surveillance methods. In routine practice, automation of the interpretation phase could point human monitors to the codes that warrant further investigation by clinical experts.

The limitations of full reliance on diagnosis codes motivate exploring automated methods of using the notes for surveillance of AEs.

We look forward to accessible methods for public health analysts to observe patterns in patient care and experience in real time, by detecting both anticipated and unanticipated signals in electronic health records that indicate potential safety concerns with medical care. Such a method could supplement existing safety surveillance methods.

## Data Availability

Original data (MIMIC-III) are available from https://mimic.physionet.org/about/mimic/ after receiving their permission.

## ACKNOWLEDGEMENTS

We thank enthusiastic support by our FDA and Booz Allen Hamilton supervisors, Department of Health and Human Services innovation programs (Ignite Accelerator and Data Science CoLab), and Alistair Johnson, DPhil, of the MIMIC-III program, Massachusetts Institute of Technology. George Plopper, PhD, Booz Allen Hamilton, provided project and consultation support. Many FDA colleagues offered ideas and feedback regarding the selection of the case of blood transfusion and the final paper. All authors had access to the data. Drs. Bright, Bright-Ponte, and Palmer are responsible for the study topic, design, and interpretation. Drs. Rankin and Bright formed the study groups. Drs. Bright and Blok are responsible for data analysis.

## Financial support

The research was done with FDA support and under contract HHSF223201510027B between FDA and Booz Allen Hamilton Inc.

## Conflicts of interest

None of the authors have other relevant financial interests.

## Disclaimer

The opinions are those of the authors and do not represent official policy of either the FDA or Booz Allen Hamilton.

## ABBREVIATIONS USED MORE THAN ONCE

95% LCL: normal approximate 95% lower confidence limits 95%
UCL: normal approximate 95% upper confidence limits
AE: adverse events
C: comparison group of admissions
EHRs: electronic health records
FDA: Food and Drug Administration
FET: Fisher’s exact test
GI: gastrointestinal
ICD9: International Statistical Classification of Diseases and Related Health Problems 9-CM
MIMIC-III: Medical Information Mart for Intensive Care III
NOS: Not otherwise specified
PTAE: potential transfusion adverse event
RD: rate difference
RR: rate ratio
T: group of transfusion admissions
TACO: transfusion associated circulatory overload
TAE: transfusion adverse event
TRALI: transfusion related acute lung injury

## Appendix to

### Bright et al, “Use of Diagnosis Codes for Blood Transfusion Adverse Events in Electronic Health Records”

**Table A1.**
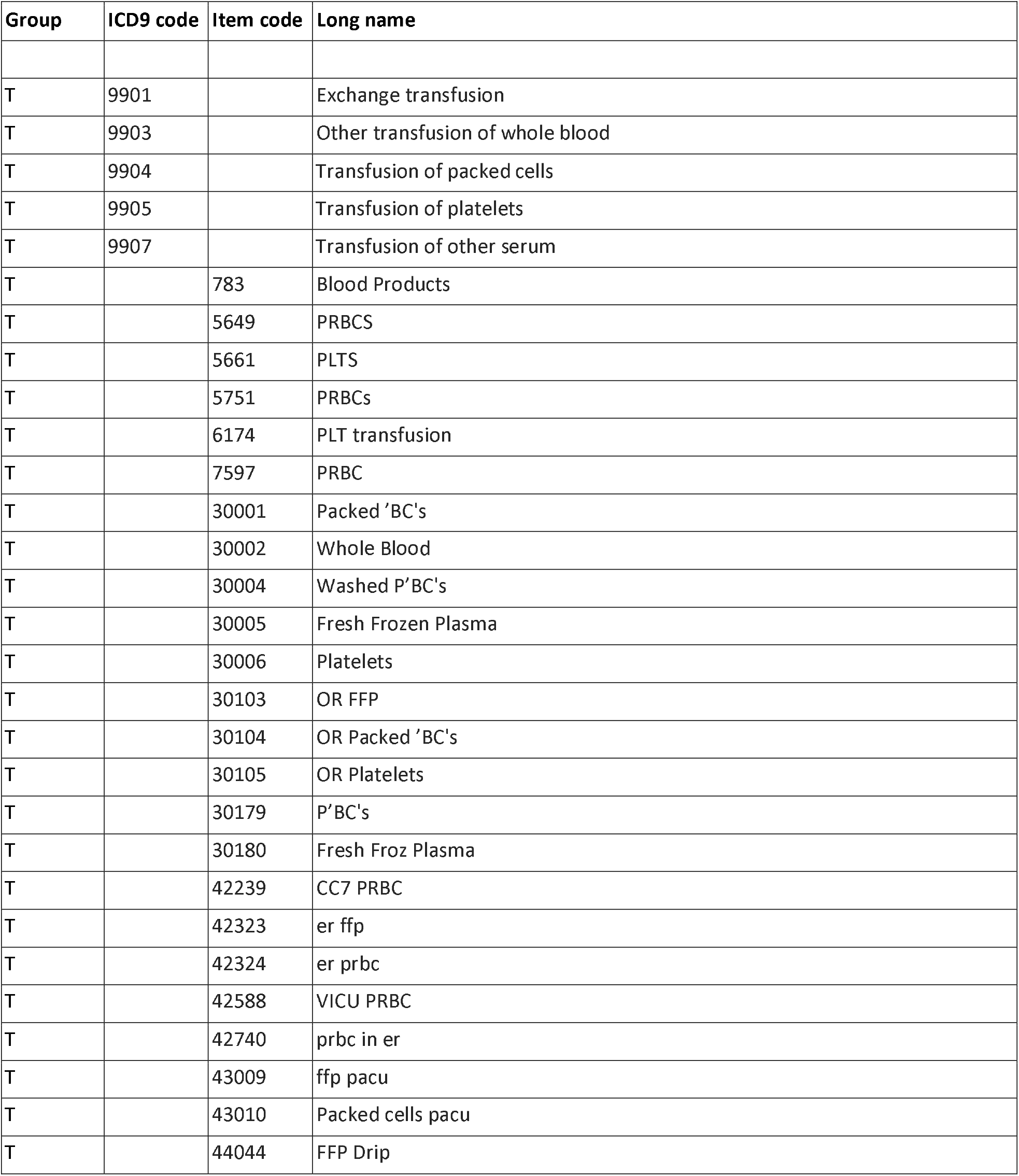
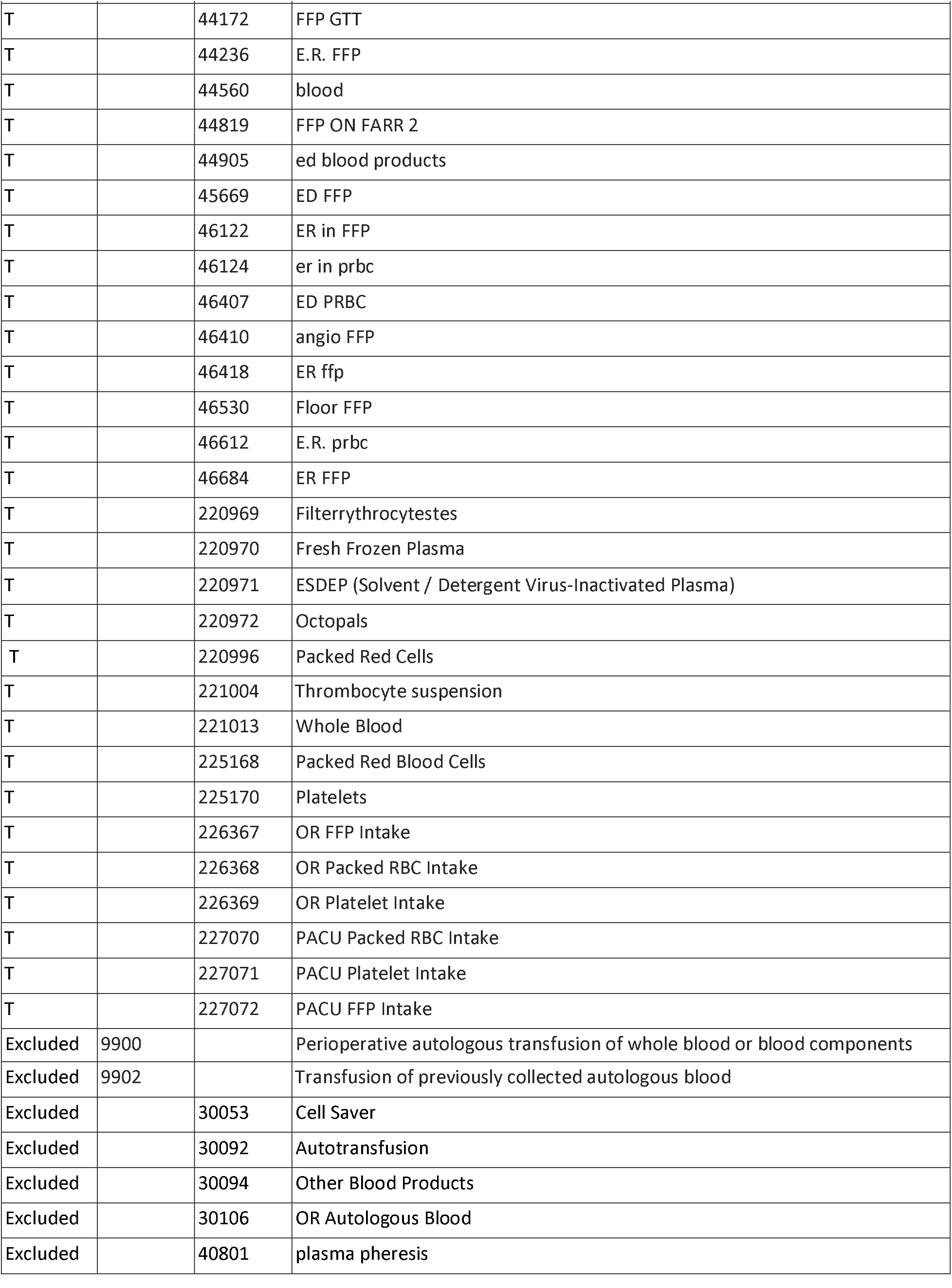
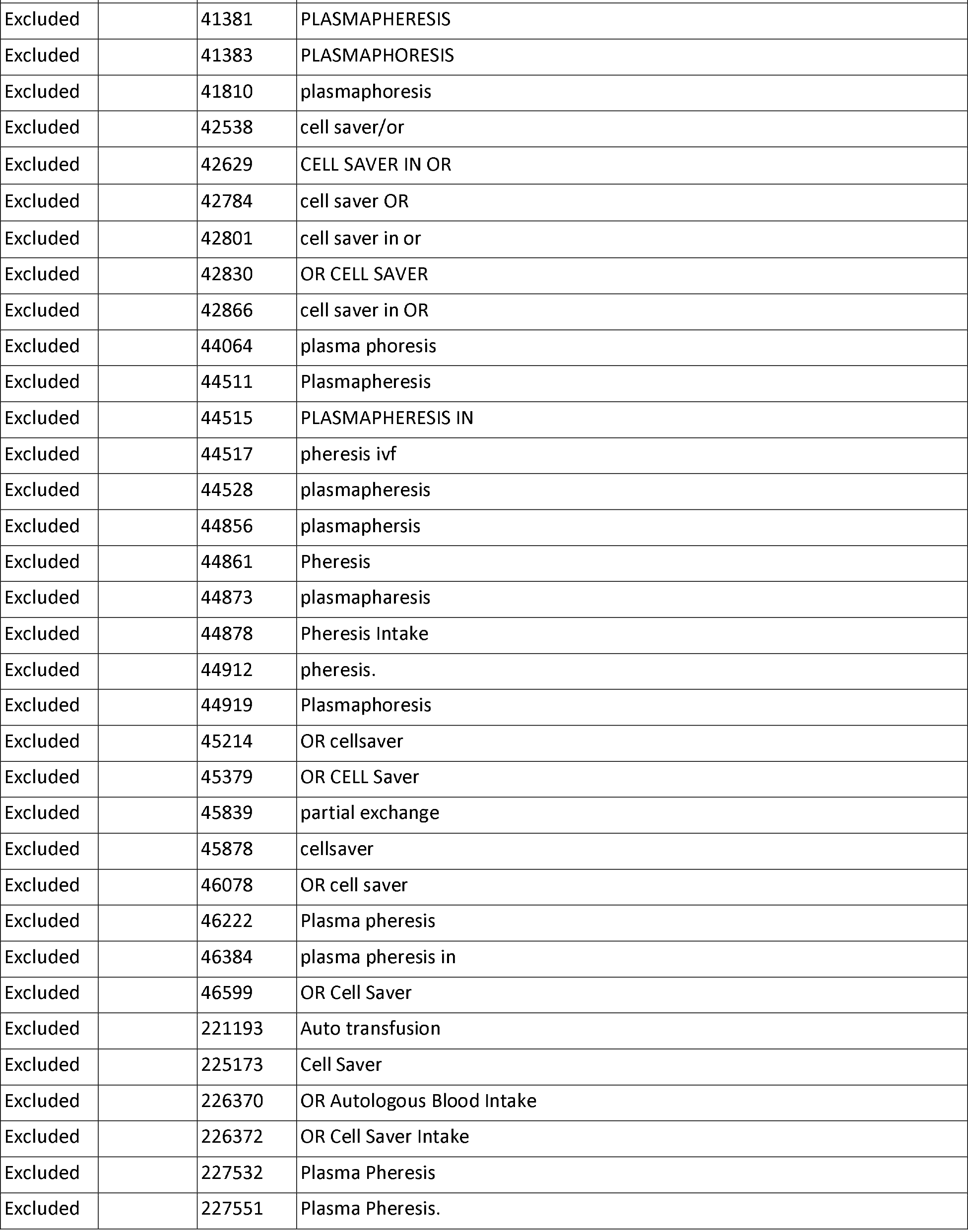
Codes for definitions of T (transfused) and Excluded groups. See main paper for methods.

## Notes

### Competing Interest Statement

The authors have declared no competing interest.

### Author Declarations

On behalf of the Food and Drug Administration Institutional Review Board (FDA IRB), Karen Ulisney, MSN, CRNP, Human Subject Protection (HSP) Executive Officer, stated the study "does not require FDA IRB review and approval because it is not research involving human subjects as defined in 45 CFR part 46 and it is not an FDA-regulated clinical investigation, as defined in 21 CFR part 56.” Patients/participants were never identified to the authors. The creators of the research dataset at the Massachusetts Institute of Technology (MIT) met the MIT Institutional Review Board requirements. This was not a clinical trial or interventional study.

